# Leveraging interindividual variability of regulatory activity refines genetic regulation of gene expression in schizophrenia

**DOI:** 10.1101/2021.10.18.21264945

**Authors:** Maris Alver, Nikolaos Lykoskoufis, Anna Ramisch, Halit Ongen, Emmanouil T. Dermitzakis

**Affiliations:** Department of Genetic Medicine and Development, University of Geneva, Geneva, Switzerland; Swiss Institute of Bioinformatics, University of Geneva, Geneva, Switzerland; Institute of Genetics and Genomics in Geneva, University of Geneva, Geneva, Switzerland; Estonian Genome Center, Institute of Genomics, University of Tartu, Tartu, Estonia

## Abstract

Schizophrenia is a polygenic psychiatric disorder with limited understanding about the mechanistic changes in gene expression regulation. To elucidate on this, we integrate interindividual variability of regulatory activity with gene expression and genotype data captured from the prefrontal cortex of 272 cases and controls. We show that regulatory element activity is structured into 10,936 and 10,376 cis-regulatory domains in cases and controls, respectively, which display distinct regulatory element coordination structures in both states. By studying the interplay among genetic variants, gene expression and cis-regulatory domains, we ascertain that changes in coordinated regulatory activity tag alterations in gene expression levels (p=8.62e-06, OR=1.60), unveil case-specific QTL effects, and identify regulatory machinery changes for genes affecting synaptic function and dendritic spine morphology in schizophrenia. Altogether, we show that accounting for coordinated regulatory activity provides a novel mechanistic approach to reduce the search space for unveiling genetically perturbed regulation of gene expression in schizophrenia.

## INTRODUCTION

Schizophrenia (SCZ) is a severe mental illness that oftentimes leads to a lifetime of chronic disability. While several lines of evidence converge on the neurodevelopmental origin for the disorder and augment the impact of both genetic and environmental factors in disease aetiology, the pathophysiology of SCZ still remains incompletely understood with little progress in novel treatment development^1,2^.

The last decades of extensive research has yielded valuable insights into the genomic and molecular underpinnings of SCZ. Large-scale genomic analyses triggered by prior studies of strong heritability estimates for SCZ (60-80%)^3,4^ have unveiled its highly polygenic architecture^5-7^. Complemented by gene expression and chromatin profiling analyses from dorsolateral prefrontal cortex (DLPFC), the SCZ risk variants have been shown to localize in functional regulatory genomic elements with approximately half of these displaying brain tissue-specific expression quantitative trait locus (eQTL) effects^5,8-10^, and to be enriched for open chromatin and evolutionary conserved regions^11,12^. Genes identified by differential expression and genome-wide association analyses (GWAS) are associated with brain developmental pathways, synaptic function, and immune response^13-15^. While immense efforts have resulted in comprehensive SCZ-specific resources for perturbed gene expression patterns and for fine-mapping and annotating discovered disease-associated genomic associations, little is known about the regulatory machinery changes and how genetic effects are propagated onto gene expression that drive molecular abnormalities in SCZ development.

Gene transcription profiles are defined by the activity of regulatory elements (REs) that overlap with open chromatin. Since the activity of REs are modulated by genetic variation, changes in chromatin accessibility result in gene expression variability and hence constitute as an intermediate phenotype for profiling eQTL effects on gene expression^16-20^. Systematic measurement of interindividual correlation between chromatin activity levels has revealed that the variability of nearby regulatory activity is structured into well-delimited sets of cis-regulatory domains (CRDs)^21^. The coordinated activity of REs within CRDs are under tight genetic control, mediate cis and trans effects of genetic variants onto gene expression and provide a higher-order structural resolution of functional regulatory associations^21^. Accounting for the three-dimensional (3D) genome organization in cis that captures concerted effect of regulatory activity could thereby facilitate a more robust signal detection for identifying disruption in regulatory function and for delineating deviations in gene expression cascades specific to disease. To build on this concept, we set out to analyse the interplay among genetic variants, coordinated regulatory activity and gene expression to characterize genetically perturbed regulatory machinery changes specific to SCZ. To this end, we integrated genome-wide genotyping data with RE activity levels (chromatin immunoprecipitation sequencing (ChIP-sequencing profiled for histone mark H3K27ac) and transcriptomic profiles (bulk RNA-sequencing) obtained from the DLPFC of SCZ cases and control subjects (dbGaP phs000979.v3.p2). At least two levels of molecular data were available for 272 individuals: 98 SCZ cases and 174 controls (188 males and 84 females, 164 African Americans and 108 Europeans; **Supplementary Fig. 1**).

## RESULTS

### Distinct regulatory element coordination in SCZ

To study the coordination of REs, we systematically measured interindividual correlation between nearby chromatin peaks. We identified 10,938 CRDs in SCZ cases, 10,376 CRDs in controls and 11,374 CRDs in the combined set (i.e., across SCZ cases and controls), regrouping 28.9% (n=40,819), 31.4% (n=44,391) and 38.4% (n=54,278) of the peaks, respectively (**Table 1**), and capturing a higher-order structural resolution of regulatory activity. The majority of the CRDs contained two REs, while some captured correlated activity among >80 REs (mean number of REs per CRD 4.7; mean CRD length 138 kb in the combined set; **Supplementary Fig. 6**). As expected, we identified a high concordance of CRDs between SCZ cases and controls (42%; **Supplementary Fig. 7**), which represent a uniform coordination of regulatory activity in DLPFC in SCZ cases and controls. However, 58% of the CRDs showed distinctive RE coordination in SCZ cases and in controls, implying notable state-dependent differences in the regulatory machinery.

**Table 1.**
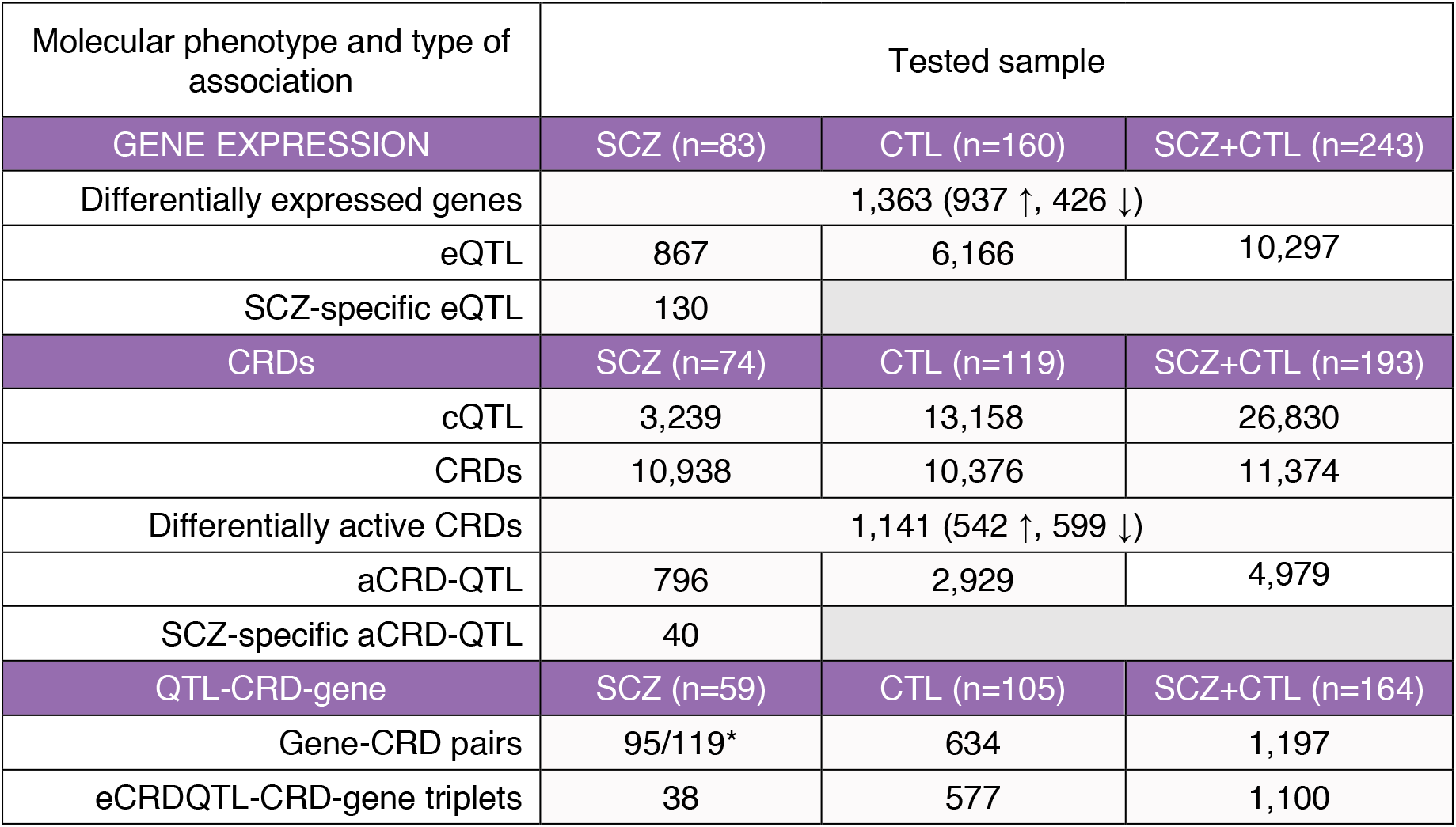
Molecular phenotype associations at FDR 5% in SCZ cases and controls. The columns indicate the molecular phenotype and type of association, and the numbers of identified associations and sample set used. *Gene-CRD associations found using CRDs identified in the combined set or CRDs identified only in SCZ cases.

Given unique RE coordination in SCZ cases and controls, we sought to investigate the mechanism for SCZ-specific CRD formation. Specifically, we asked whether the chromatin peaks within SCZ-specific CRDs were differentially active or whether these had larger variance in activity in controls compared to SCZ cases. To this end, we considered CRDs in SCZ cases composed of peaks not part of any CRD in controls and compared single peak activities and mean correlation estimates among peaks per CRD between the two groups. We discovered that 53% of the peaks within SCZ-specific CRDs (3,540 peaks in 2,212 CRDs) were differentially active in SCZ cases at false discovery rate (FDR) 5% (**Supplementary Fig. 8a**). While the majority of the peaks (71%) showed lower activity in SCZ cases (**Fig 1a**), only a third of SCZ-specific CRDs had all underlying peaks differentially active between SCZ cases and controls (**Supplementary Fig. 8b**), implying that the regulatory activity originating from those genomic regions likely results in inhibition of downstream molecular cascades. The peaks of SCZ-specific CRDs displayed significantly higher mean correlation in SCZ cases compared to controls (Mann-Whitney U test p = 5.02e-47; **Fig. 1bc**), indicating that changes in the 3D structure of the genome, rather than differential activity, were responsible for SCZ-specific CRD formation. At FDR 5%, we identified eleven SCZ-specific CRDs to be associated with the expression of proximal genes, for example *POU3F1* (also known as *OCT6*, transcriptional repressor for myelin-specific genes^22^), *KIF5A* (neuronal-specific vesicular transporter^23^), *NECAB1* (Ca^2+^-binding in neurons^24^), and *PDCD1LG2* (immune checkpoint receptor ligand^25^) (**Supplementary Table 1)**. These associations exemplify coordinated regulatory changes specific to the disease state that affect or are affected by gene expression perturbations.

**Fig. 1.**
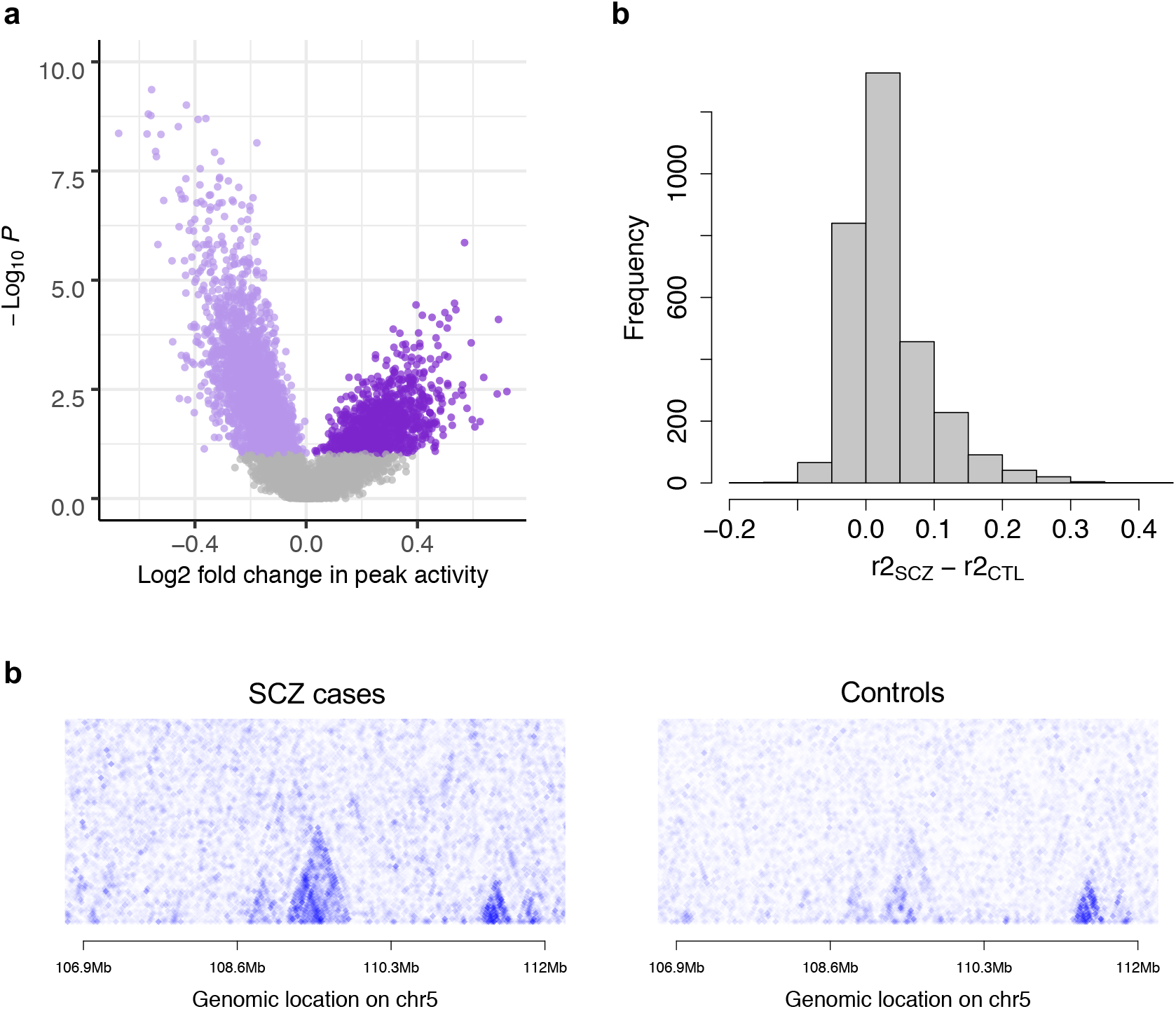
Features of SCZ-specific CRDs. (a) Difference in medians per peak activity between SCZ cases and controls as a function of the strength of association given in -log10 p-values; purple dots denote peaks that are differentially active between SCZ cases and controls at FDR 5% (3,450 peaks) with light purple and dark purple indicating lower and higher median activity, respectively, in SCZ cases compared to controls. (b) Comparison of differences in per CRD peak activity correlation estimates between SCZ cases and controls for SCZ-specific CRDs (3,078 CRDs). (c) Example region of a correlation structure between 250 peaks on chromosome 5 in SCZ cases and controls, revealing a well-delimited SCZ-specific CRD that is composed of 5 regulatory elements.

### Changes in CRD activity track alternations in gene expression in SCZ

Having identified several distinct RE coordination structures in DLPDC in SCZ cases that were absent in controls, we focused next on CRDs that had the same structure of RE coordination across SCZ cases and controls (i.e., CRDs identified in the combined set). We set out to determine differences in regulatory activity between SCZ cases and controls and investigated their relation to genes that were differentially expressed (DEGs) between the two groups. At FDR 5%, we identified 1,141 CRDs (599 lower activity, 542 higher activity) and 1,363 genes (937 up-regulated, 426 down-regulated) to be differentially active and expressed in SCZ cases, respectively (**Table 1, Supplementary Fig. 9, Supplementary Table 2, Supplementary Table 3**). The differences in effect size for CRDs were subtle, reflecting a narrow variability range in regulatory activity (**Supplementary Fig. 9a**). The determined DEGs were in concordance with those previously reported in SCZ pathogenesis (**Supplementary Fig. 10**)^9,26^ and were significantly enriched for gene ontology (GO) terms related to sex-hormone and interferon-*γ*-mediated signalling, glucocorticoid receptor and glutamate receptor binding, axonogenesis and synapse assembly (**Supplementary Table 4**). The DEGs were significantly enriched for differentially active CRDs (Fisher’s exact test p = 8.62e-06, odds ratio 1.60) (**Fig. 2a**) with the majority of the genes (86%) showing the same direction of effect as the CRD in which the gene transcription start site (TSS) lied, indicating that deviations in gene expression track alterations in the regulatory machinery.

**Fig. 2.**
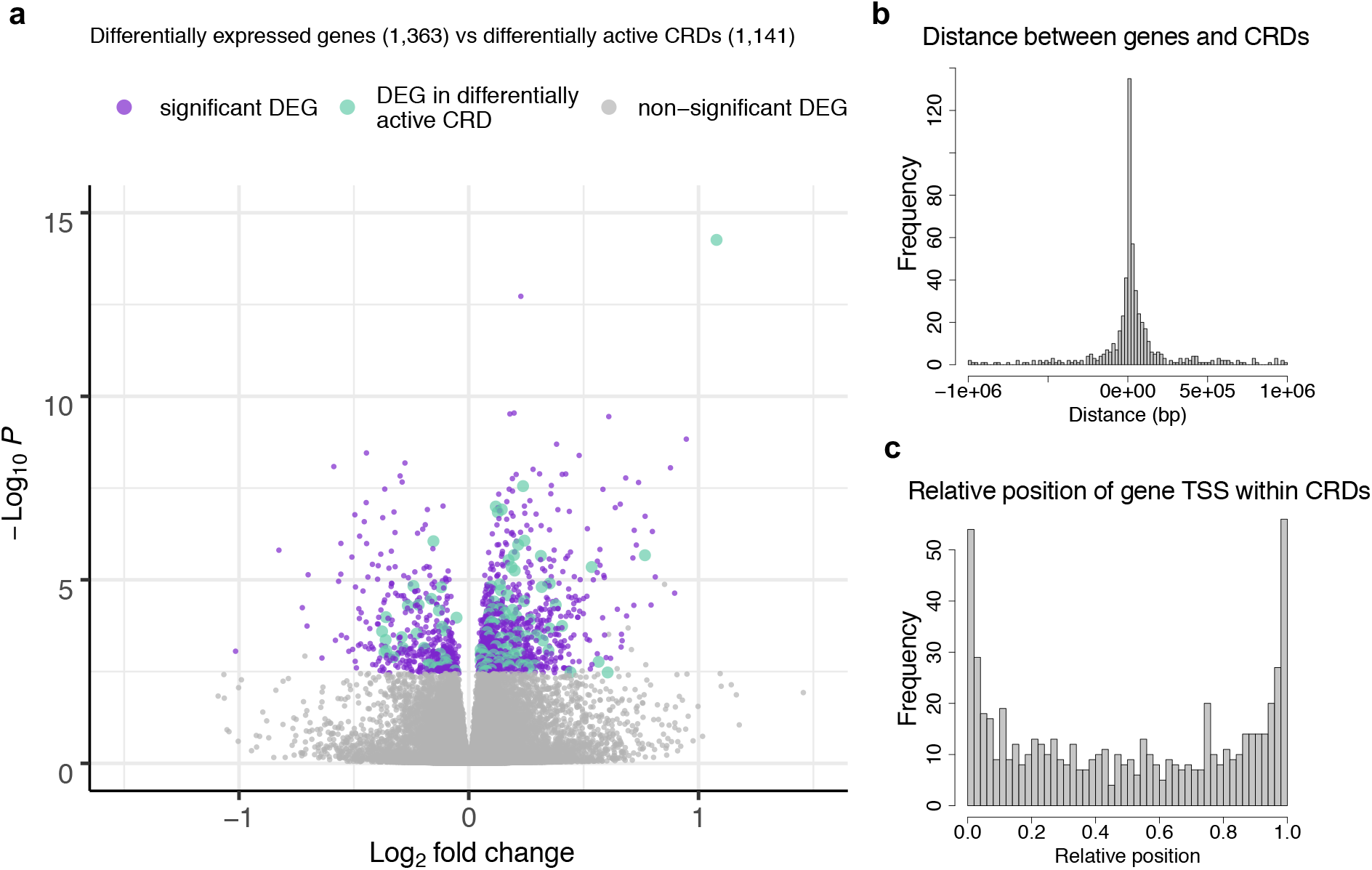
Association between CRDs and genes. (a) TSSs of differentially expressed genes (DEG) are localized within differentially active CRDs significantly more often than expected by chance (Fischer’s exact test p = 8.72 × 10^−6^, odds ratio 1.60); coloured dots denote DEGs identified at FDR 5%: purple dots mark DEGs, turquoise dots mark DEGs with TSS within differentially active CRD. (b) Distribution of gene-to-CRD distances for genes localizing outside the associated CRD boundary (545 gene-CRD associations). (c) Distribution of the relative position of gene TSS to the boundary of an associated CRD (652 gene-CRD associations).

In addition to ascertaining that differentially active CRDs localized within the genomic proximity of DEGs, we additionally tested for direct association between gene expression and CRD activity (i.e., CRDs identified in the combined set). At FDR 5%, we identified in cis 95, 634 and 1,197 CRD-gene associations in SCZ cases, in controls, and in the combined set, respectively (**Table 1, Supplementary Fig. 11, Supplementary Table 6**). The majority of the genes were associated with a single CRD and the majority of the CRDs with a single gene with only a handful of CRDs being associated with up to ten different genes (**Supplementary Fig. 12**). Most gene TSSs clustered at CRD boundaries (**Fig. 2bc**), corroborating the proximal role of coordinated regulatory activity in gene transcription.

### Genetic regulation of CRD activity and gene expression in SCZ

We next sought to study the genetic regulation of CRD activity and gene expression, search for SCZ-specific QTL effects and interrogate whether QTL effects colocalize with SCZ risk variants. At 5% FDR and in cis, we discovered 796 and 2,929 functionally independent CRD activity QTLs (aCRD-QTLs), and 867 and 6,166 functionally independent eQTLs in SCZ cases and controls, respectively (**Table 1, Supplementary Table 6, Supplementary Table 7, Supplementary Fig. 13**). The strength of the association was correlated with the genomic distance from the molecular phenotype (**Supplementary Fig. 14**). While almost all SCZ-identified QTL effects replicated in controls (**Supplementary Fig. 15**), 5% of aCRD-QTLs (n=40) and 15% of eQTLs (n=130) showed SCZ-specificity, i.e., these affected CRD activity or gene expression only in SCZ cases or displayed significant change in effect size compared to controls (**Table 1, Fig. 3ab**). The SCZ-specific genotype-dependent variability in CRD activity and in gene expression imply context-dependent and pathway-activated gain in regulatory capacity. Results of gene enrichment analysis for genes associated with SCZ-specific eQTLs conform with posed hypotheses linking dysregulation of retinoid binding and adenosine deaminase activity with SCZ^27,28^ (**Supplementary Table 8**). Colocalization analyses for SCZ risk variants with aCRD-QTLs and with eQTLs showed modest yet proportionally similar enrichment for shared functional effects (1.6% for SCZ-identified aCRD-QTLs and 1.8% for SCZ-identified eQTLs). Interestingly, the aCRD-QTLs colocalized with different GWAS variants compared to eQTLs that shared a functional effect with SCZ risk variants (**Supplementary Table 9**).

**Fig. 3.**
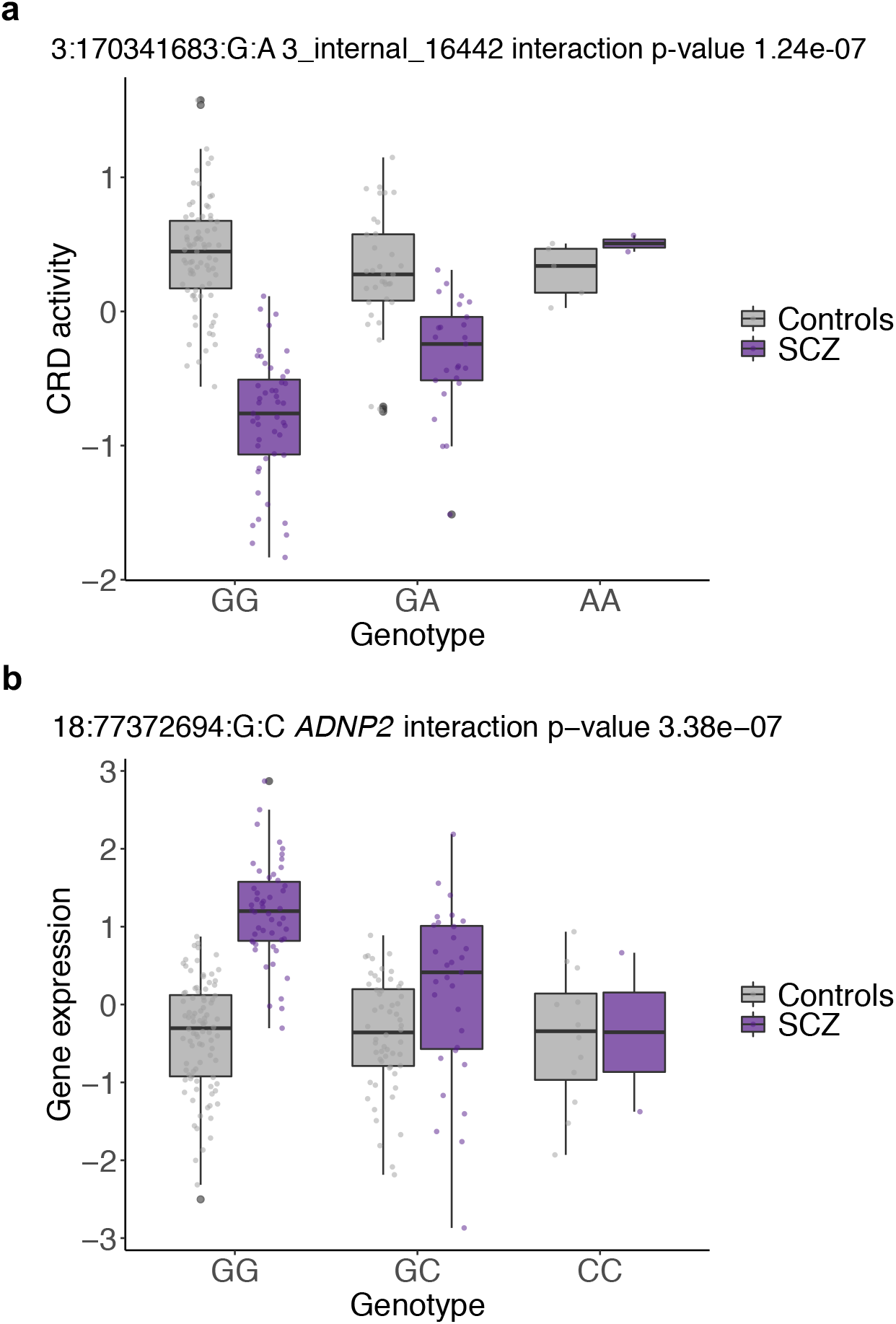
Example of a SCZ-specific (a) aCRD-QTL and (b) eQTL effect. Genotype-dependent effect on CRD activity and gene expression identified only in SCZ cases.

To assess common genetic regulation of coordinated regulatory activity and gene expression, we investigated the association of aCRD-QTL and eQTL effects on the other molecular phenotype. Specifically, we correlated aCRD-QTLs with gene expression and eQTLs with CRD activity over gene-CRD associations detected across all samples at nominal significance. We identified that up to 46% of the eQTL and aCRD-QTL variants had an effect on CRD activity and gene expression, respectively (**Supplementary Fig. 16**). The considerable overlap between aCRD-QTL and eQTL effects for relevant CRD-gene pairs corroborates the functional interplay among genetic variants, CRDs and genes.

### Refining eQTL perturbations reveals regulatory machinery changes specific to SCZ

Given the established interplay among genetic variants, genes and CRDs, we interrogated the functional directionality between them. We reasoned that the effect of a genetic variant on gene expression could either be mediated by or propagated to the changes in coordinated RE activity and that deviations in the regulatory machinery in SCZ cases compared to controls would imply molecular dysregulation specific to disease. To test this, we considered the previously discovered 1,197 CRD-gene pairs ascertained across SCZ cases and controls at FDR 5% and identified the same genetic variant (eCRD-QTL) that affected both molecular phenotypes and by that determined eCRDQTL-CRD-gene triplets for causal inference (**Methods**). Using Bayesian Networks (BN), we tested three relationship patterns: i) causal model in which the genetic variant affects first the CRD activity which then regulates the gene expression, ii) reactive model in which the genetic variant affects the gene expression which modulates the CRD activity, and iii) independent model in which the genetic variant affects the gene and the CRD independently; and studied these relationships separately in SCZ cases (n=59) and controls (n=105) (**Supplementary Fig. 17**). We discovered that at FDR 5%, 91.9% of the CRD-gene pairs had a cis-QTL effect (n=1,100; **Table 1**), indicating that the simultaneous change in CRD activity and gene expression was affected by the same nearby genetic driver. We observed more causal models in controls than in SCZ cases (**Supplementary Fig. 18ab**), which were likely driven by smaller SCZ sample size as reflected by the distribution of the probabilities for the most likely model for each triplet (**Supplementary Fig. 18cd**). The probability of the causal model increased the further the gene TSS was from the eCRD-QTL in both SCZ cases and controls (**Supplementary Fig. 18ef**), denoting the role of CRDs mediating the genetic effect onto distal genes.

To study the proportion of differential regulatory mechanisms between SCZ cases and controls and ascertain which molecular functions were affected by these changes, we first estimated the accuracy for BN results using bootstrapping to provide confidence for retrieved probabilities and next carried out gene enrichment analyses for genes associated with different regulatory mechanisms. The accuracy for inferring the most likely causal relationship for triplets was lower in SCZ cases (mean accuracy estimation 69.0%, sd = 16.6) than in controls (mean accuracy estimation 75.0%, sd = 18.0) (**Supplementary Fig. 19ab**). To exclude ambiguous signals, we proceeded with triplets that surpassed the accuracy estimation of 55% (748 triplets, 68% of studied triplets; **Methods, Supplementary Fig. 19c, Supplementary Table 10**). While two-thirds of the triplets displayed the same regulatory mechanism in SCZ cases and controls (**Fig. 4a)**, one-third of studied triplets showed a change in directional effect from QTL variant onto molecular phenotype (**Fig. 4b, Supplementary Fig. 19d**). These deviations in regulatory mechanism in SCZ reflect gain or loss in the regulatory capacity that could either be driven by context-dependent or genetically predisposed developmental derailment of gene expression, or affected by external stimuli (e.g., treatment). The genes associated with change-associated triplets were significantly enriched for GO terms related to small GTPase binding, filopodium assembly and cellular lipid catabolic process (**Supplementary Table 11**), highlighting alterations in the regulatory machinery for gene expression affecting synaptic function and plasticity, and dendritic spine morphology in SCZ.

**Fig. 4.**
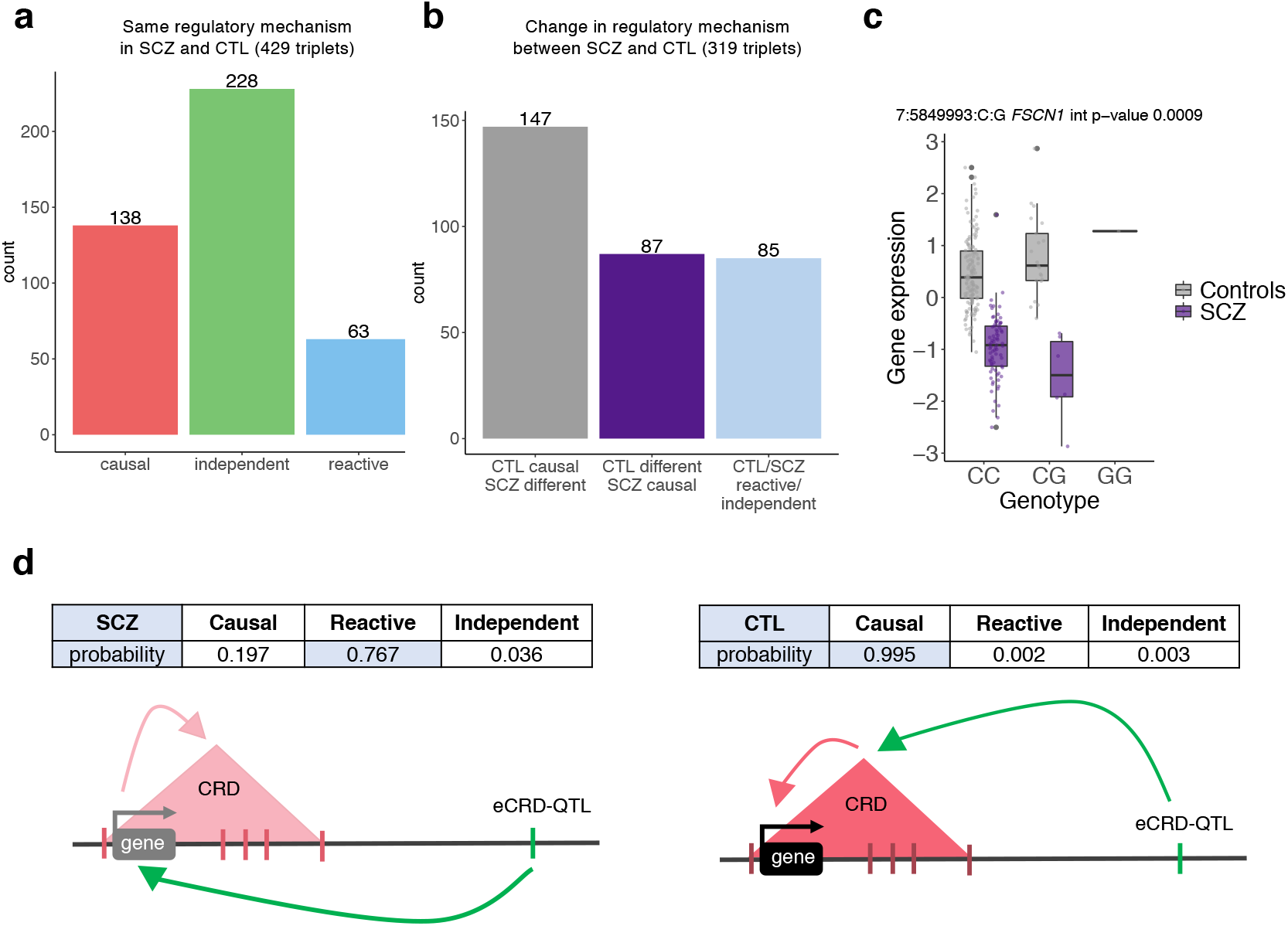
Regulatory mechanisms for eCRDQTL-CRD-gene triplets in SCZ cases and controls.: Comparison of the direction of effect from eCRD-QTL onto gene expression and CRD activity for tested triplets between SCZ cases and controls: (a) triplet count for models showing the same regulatory mechanism in SCZ cases and controls, and (b) triplet count for models showing a change in the regulatory mechanism between SCZ cases and controls (term “different” indicates either reactive or independent model; light blue bar indicates triplets, for which the causal model (i.e., mediation via CRD for QTL effect) was not identified in SCZ cases nor in controls). (c) Genotype-dependent effect for eCRD-QTL 7:5849993:C:G on *FSCN1* expression. (d) Distinct regulatory mechanism of genetic regulation on gene expression for SCZ cases and controls for a triplet consisting of an eCRD-QTL 7:584993:C:G, gene *FSCN1* and a CRD composed of 5 REs on chr7:5623132-5705414; the probabilities based on Bayesian Networks for each tested model is given above schematics; shading of the colour for the gene and for the CRD indicates strength in expression and activity, respectively.

Perturbation in the regulatory mechanism of gene expression in SCZ is exemplified by a triplet consisting of an eCRD-QTL 7:5849993:C:G associated to gene *FSCN1* and to a CRD composed of 5 REs (chr7:5623132-5705414) (**Supplementary Fig. 20**). FSCN1 is an actin-binding protein that is required for the formation of actin-based cellular protrusions and affects dendritic spine morphology^29-32^. Based on BN, the genetic variant affected first the activity of the CRD and then the expression of the gene in controls (causal model probability 0.99), whereas in SCZ cases the change in CRD activity was a reaction to gene expression (reactive model probability 0.77), indicating that the eCRD-QTL effect on *FSCN1* expression was not mediated via the associated CRD activity in SCZ cases as seen for controls (**Fig. 4d**). This was further supported by significant downregulation of *FSCN1* expression and CRD activity in SCZ cases compared to controls (FDR 5% p-value 0.02 and 0.005, respectively; **Supplementary Fig. 20ab, Supplementary Table 2, Supplementary Table 3**). Moreover, we identified an opposite direction of eCRD-QTL effect on *FSCN1* expression in SCZ cases compared to controls (**Fig. 4c**), but not on CRD activity (**Supplementary Fig. 20c**). Interestingly, this association was identified only in individuals with African American ancestry (**Supplementary Fig. 20de**) as the genetic variant was completely monomorphic in HBCC Europeans. The MAF spectrum of 12% in HBCC African Americans and 0% in HBCC Europeans is in concordance with population frequencies estimated in larger datasets (MAF 13% in Africans/African Americans and 0.4% in non-Finnish Europeans)^33^. These results indicate that the downregulation of *FSCN1* expression in SCZ cases was driven by a different intermediatory regulatory mechanism that deviated from one seen in controls and represents a dysregulated step within an abnormal molecular cascade affecting dendritic spine morphology in SCZ.

## DISCUSSION

Deciphering regulatory mechanisms of gene expression that reflect molecular perturbation in SCZ are under extensive scrutiny yet are hindered by the complexity of the SCZ phenotype and scarcity of relevant molecular data. Studying regulatory activity that tracks changes in gene expression requires a higher order analysis approach for signal discernment due to narrow variability range in regulatory activity and extensive multiple testing burden^12^. Here we show that taking account of interindividual correlation between regulatory activity allows to refine changes in gene expression specific to disease, asserting that disease manifestation stems from dysregulated gene expression cascades that are steered by and propagated to the concerted action of REs. Interrogation of common genetic regulation of gene expression and CRD activity corroborated that correlated changes in gene expression and CRD activity are affected by the same genetic driver. Our results agree with findings showing considerable overlap between QTL effects on chromatin accessibility and gene expression^12^, that a single genetic variant drives the association between multiple chromatin peaks and a single gene^20^, and on the convergence of deviations detected in different molecular layers as seen for gene expression and methylation in SCZ^34^. Applying causal inference to study the causal relationships among genetic variants, genes and CRDs revealed regulatory machinery changes affecting synaptic function and dendritic spine morphology in SCZ which are in line with established molecular abnormalities identified for the disorder^35-39^. The deviations in regulatory mechanism reflect gain or loss in the regulatory capacity that could either stem from genetic predisposition, are acquired in disease progression or result from chronic pharmacology. Clear discernment of the proposed origins of effect was hampered due to small sample size and unavailability of relevant data yet allowed to draw the following conclusions.

First, while we found modest colocalization for detected QTLs with SCZ-predisposing genetic variants, the results reflect direct correlation between sample size and QTL signal detection^40^ and hence are in proportion to colocalization signals ascertained in previous findings^9,12,26,41^. Previous studies have highlighted the concordance of SCZ heritability enrichment for open chromatin regions in fetal and in SCZ DLPFC samples and the stability of methylation and expression feature deviations in fetal brain development that persist into adulthood for those affected by the disorder^9,12,13,42^, implying that a considerable proportion of signals detected in the current analysis do reflect brain development derailment due to genetic predisposition for SCZ. Second, while molecular data used in this study was extracted from bulk tissue, consistent comparison with signals identified in control samples provides confidence that the identified deviations in SCZ cases captured the most notable disease-specific molecular abnormalities in DLPFC. This is further supported by investigations revealing that DLPFC transcriptomic profiles are generally biased toward neuronal populations^34^ and that SCZ risk variants are overrepresented in neuronal vs non-neuronal open chromatin regions^43^. Third, studying QTL effects on gene expression and CRD activity separately in SCZ cases and controls allowed to discriminate context-dependent genetic effects on both molecular phenotypes, indicating gain in regulatory capacity that translated into gene expression and coordinated regulatory activity variability only in SCZ cases or showed significantly different or even discordant effect direction, as seen for *FSCN1* gene, between the two groups. Fourth, a previous study that showed concordant DEG signals with those found in the current analysis, identified that differential gene expression in SCZ was not driven by antipsychotic intervention^26^, ensuring that treatment effect was not the main trigger for DEG results between SCZ cases and controls in the current analysis. Lastly, inclusion of individuals of European and African American ancestry augmented signal identification and corroborates that the genetic basis of SCZ and its biology are broadly shared across populations^7,44,45^.

Altogether, we have outlined that leveraging higher-order structural resolution of regulatory activity allows to reduce the search space for unveiling genetically perturbed regulation of gene expression specific to SCZ. We anticipate that cell-type specific gene expression and open chromatin exposure profiles in larger sample sets would allow better delimitation of CRD chromatin peak content, facilitate the identification of trans-regulatory hubs across different chromosomes as well as enhance more robust detection of origin effect for gene expression deviation, thereby increasing our understanding of perturbed functional pathways underlying schizophrenia and for prioritizing targets for experimental investigation and novel treatment development.

## METHODS

### Molecular and phenotype data

Molecular and phenotype data for the Human Brain Collection Core (HBCC) was accessed through dbGaP (study accession phs000979.v3.p2; request #88083-1 approved by NIH on January 31^st^, 2020). All patients met DSM-IV criteria for a lifetime Axis I diagnosis of psychiatric disorders including schizophrenia or schizoaffective disorder. Controls had no history of psychiatric diagnoses or addictions. At least two levels of molecular data (i.e., whole-genome genotype, RNA-sequencing or ChIP-sequencing data) were available for 272 individuals. For genotype data, we determined the intersect of single nucleotide variant (SNV) content across three Illumina genotyping arrays (HumanHap650Y, Human1M-Duov3 and HumanOmni5M-Quad) after filtering the SNVs using standard procedure with PLINK v2.0^46^ and imputed the derived genotype matrix using the Haplotype Reference Consortium reference panel^47^. Next, we applied post-imputation quality control filters by European and African American ancestral group separately and considered the union of filtered SNVs retrieved in both ancestry sets for the final SNV set. This yielded 8,245,179 biallelic SNVs in 272 individuals. Sequence data was mapped onto the human genome (hg19) with either BWA-MEM v0.7.16^48^ for ChIP-sequencing data or STAR^49^ for RNA-sequencing data. Gene expression was quantified using QTLtools^50^ and filtered for protein-coding and lincRNA genes and for the union quantifications detected in ≥ 50% in SCZ cases and in ≥ 50% in controls. This yielded 21,988 genes for 243 individuals. ChIP-sequencing peak calling and quantification was carried out with HOMER v14.11.1^51^. We first determined ChIP-sequencing peak coordinates across SCZ cases and controls to get a population scale call set of ChIP-sequencing peaks and then quantified the peaks for each individual according to the identified peak coordinates. This yielded 141,219 ChIP-sequencing peaks for 193 individuals. To account for confounding factors in gene expression and ChIP-sequencing peak data, we regressed out ancestry (captured by principal component (PC) analysis on the genotype data), and technical variables (captured by PC analysis on the molecular phenotype data). For the latter, we used the number of PCs that maximized the number of QTLs discovery. Both gene expression and ChIP-sequencing data were normalized such that these matched a normal distribution with mean 0 and standard deviation 1.

### Cis-regulatory domain calling and quantification

For CRD calling, we used the pipeline developed in Delaneau et al. 2019^21^. First, we built a correlation map from chromatin data by systematically measuring interindividual correlation (i.e., Pearson correlation coefficient) between all possible pairs of ChIP-sequencing peak quantifications located on the same chromosome (within a 250-peak sliding window). Next, we applied hierarchical clustering on the data on a per chromosome basis to get a binary tree that regroups chromatin peaks for each chromosome depending on the correlation levels they exhibited. We relied on three empirical criteria for CRD calling: (i) overall correlation that requires the mean level of correlation within a CRD to be at least twice the background, (ii) edge correlation that requires the mean level of correlation of the peaks at the CRD boundaries to be at least twice the background, and (iii) a requirement that the CRD covers at least two nonoverlapping regulatory elements. We quantified CRD activity on a per-individual basis by enumerating all ChIP-sequencing peaks per CRD and taking the mean of all single peak quantifications per individual to retrieve a single quantification value for each individual. CRDs called in SCZ cases were used for characterizing SCZ-specific CRDs. CRDs identified in the combined set were used for the rest of the downstream analyses.

### CRD structure analysis

For determining CRD sharing between SCZ cases and controls, we compared ChIP-sequencing peak correlation maps between SCZ cases and controls and called a CRD shared if ≥ 50% of the peaks overlapped. To assess the features of SCZ-specific CRDs, we considered only CRDs in SCZ cases composed of peaks not regrouping into any CRD in controls. For peak activity estimation, we used ChIP-sequencing peak quantifications uncorrected for covariates and applied a Mann-Whitney U test per peak activity between SCZ cases and controls. Significant differences between SCZ cases and controls were determined at FDR 5% using *R/qvalue* package^52^. For estimating peak activity correlation per SCZ-specific CRD between SCZ cases and controls, we used ChIP-sequencing peak quantifications corrected for biological and technical covariates, calculated the mean Pearson correlation estimate between peak activities per CRD and used Mann-Whitney U test for comparing these correlation estimates between SCZ cases and controls.

### Differential CRD activity and differential gene expression analysis

Differential CRD activity analysis and differential gene expression analysis were carried out using DESeq2^55^. Significant associations were determined at FDR 5%^52^. For differential CRD activity analysis, we used unnormalized ChiP-sequencing peak read counts obtained with HOMER^51^ and summed these up per CRD using the correlation map identified in the combined set. For differential gene expression analysis, we used RNA-sequencing read counts. To identify covariates for correction, we carried out an association testing i) between all available biological and technical covariates and disease status (SCZ/CTL) (Mann-Whitney U test), and ii) between all available biological and technical covariates and individual ChIP-sequencing peak activity or gene expression quantifications (linear regression) and calculated π1 estimate^52^ to identify the proportion of true associations. R*/clusterProfiler* package^56^ was used for gene set enrichment analysis.

### Mapping molecular quantitative trait loci (QTLs)

QTL mapping was carried out using the standard procedure implemented in the QTLtools software package^50^. Specifically, we performed 1,000 permutations to correct for the number of genetic variants being tested in cis per molecular phenotype (+/-1 Mb window) and corrected for the number of molecular phenotypes being tested genome-wide using false discovery rate (FDR)^52^. To identify multiple QTLs with independent effects on a molecular phenotype, we used the conditional analysis approach based on a forward-backward scan implemented in QTLtools^50^. For SCZ-specific QTL discovery, we considered QTL effects identified in SCZ cases and for each variant-phenotype pair ran a linear regression including genotype, disease status (SCZ/CTL), and covariates, and tested for significance of a genotype * disease status interaction on molecular phenotype (gene expression or CRD activity). This was followed with FDR 5% correction for the number of QTLs tested. We assessed the likelihood of a shared functional effect between SCZ risk variants from four GWAS studies^5-7,45^ and SCZ-identified QTLs using regulatory trait concordance (RTC)^53,54^.

### CRD and gene association

We used QTLtools cis permuation pass^50^ to identify CRDs associated with a gene in a +/-1 Mb window from a transcription start site of a gene. We performed these analyses i) to identify genes associated with SCZ-specific CRDs using CRDs identified in SCZ cases, and ii) to capture comparable associations for SCZ cases and controls using CRDs detected in the combined set. We tested QTL effects for association with the other molecular phenotype (i.e., aCRD-QTLs with gene expression eQTLs with CRD activity) via CRD-gene associations detected across all samples at nominal significance.

### Causal inference

To quantify gene-CRD pairs identified as significant at FDR 5% across SCZ cases and controls, we used PC analysis-based dimensionality reduction. For each gene-CRD pair, we aggregated gene expression with CRD activity and used the coordinates on PC1 as new pseudo-phenotypes for QTL mapping in a cis window. This effectively gave us eCRDQTL-CRD-gene triplets consisting of a genetic variant, a CRD and a gene, all associated with each other.

We applied a Bayesian Network approach to infer the most likely causal relationship for eCRDQTL-CRD-gene triplets common to SCZ cases and controls and conducted the analyses separately in SCZ cases and in controls. This approach allowed to estimate the most likely network from which the observed data originates by calculating the posterior probabilities for the three possible causal models^57^: i) causal model in which the genetic variant affects first the CRD and then the gene, ii) reactive model in which the genetic variant affects the gene and then the CRD, iii) independent model in which the genetic variant affects the gene and the CRD independently.

To provide confidence for retrieved probabilities, we carried out 100 bootstrapping runs for each tested triplet separately for SCZ cases and controls using sampling with replacement. We estimated how many times the most probable model across bootstrapping runs for each triplet was the same as in the original Bayesian Network results and filtered out all triplets that fell below a confidence threshold of 55%: this corresponds to the lower quartile value in SCZ cases. *R/clusterProfiler* package^56^ was used for gene set enrichment analysis for genes that belonged to triplets showing directional change from eCRD-QTL onto gene expression/CRD activity between SCZ cases and controls. We considered two scenarios: i) causal model in controls, but reactive/independent in SCZ cases; ii) reactive/independent in controls, but causal in SCZ cases.

## Data Availability

All data produced in the present study are available upon reasonable request to the authors.

## ACKNOWLEDGEMENTS

This research was supported by the Intramural Research Program of the NIMH (NCT00001260, 900142), the Estonian Research Council ETAg grant no PUTJD901 (to M.A.) and the National Centre of Competence in Research (NCCR) grant “Synapsy – The Synaptic Bases of Mental Diseases” (no 51NF40-158776) from the Swiss National Science Foundation. The funders had no role in study design, data collection and analysis, decision to publish, or preparation of the manuscript.

## Notes

### Competing Interest Statement

Emmanouil Dermitzakis is currently an employee of GSK. The work presented in this manuscript was performed before he joined GSK.

### Author Declarations

Phenotype and molecular data for the HBCC cohort are deposited at dbGaP (study accession phs000979.v3.p2; https://www.ncbi.nlm.nih.gov/projects/gap/cgi-bin/study.cgi?study_id=phs000979.v3.p2). Genotype data for 1000 Genomes Project are deposited at https://www.internationalgenome.org/data-portal/data-collection.

