## Supplementary Material for "Leveraging interindividual variability of regulatory activity refines genetic regulation of gene expression in schizophrenia"

This PDF file includes:
Materials and Methods
Figures S1 to S20
References

DATA PREPARATION

General overview

Molecular data for the Human Brain Collection Core (HBCC) within the Division of Intramural Research Programs (DIRP) at the National Institute of Mental Health (NIMH) was accessed through dbGaP (study accession phs000979.v3.p2; request #88083-1 approved by NIH on January 31st, 2020; https://www.ncbi.nlm.nih.gov/projects/gap/cgi-bin/study.cgi?study_id=phs000979.v3.p2).

Briefly, post-mortem dorsolateral prefrontal cortex (DLPFC) brain tissues were obtained under the protocols approved by the CNS IRB with the permission of the next-of-kin through the Offices of the Chief Medical Examiners in the District of Columbia, Northern Virginia, and Central Virginia, and from the University of Maryland Brain and Tissue Bank and the Stanley Medical Research Institute. Clinical characterization, neuropathological screening, toxicological analyses, and the dissections of the brain region were performed as described previously[1]. All patients met DSM-IV criteria for a lifetime Axis I diagnosis of psychiatric disorders including schizophrenia or schizoaffective disorder. Controls had no history of psychiatric diagnoses or addictions. While the HBCC cohort consists of individuals with different ancestral background, we opted for using samples from the two largest self-reported ancestral groups: African American and European.

Phenotype data

At least two levels of molecular data (i.e., whole-genome genotype, RNA-sequencing or ChIP-sequencing data) were available for 272 individuals: 98 SCZ cases and 174 controls, 188 males and 84 females, 164 African Americans and 108 Europeans; controls <16 years of age were excluded (**Supplementary Fig. 1**).

Genotype data

Subjects were genotyped with three Illumina whole-genome genotyping arrays: HumanHap650Y, Human1M-Duov3 and HumanOmni5M-Quad. Raw .idat files were downloaded from dbGaP and converted to .gtc and .vcf files using Illumina Array Analysis Platform Genotyping Command Line Interface (iaap-cli and gtc2vcf (webpage: https://github.com/freeseek/gtc2vcf), respectively. Array-based quality control was carried out with PLINK v2.0[2] (webpage: https://www.cog-genomics.org/plink/2.0/) using the following criteria: i) exclusion of individuals with genotype call rate <95%; and exclusion of single nucleotide variants (SNVs) with call rate <95%, Hardy-Weinberg equation (HWE) <10e-5, minor allele frequency (MAF) <0.01, and with ambiguous genotypes (AT and GC SNVs); ii) confirmation of the match for genotype and phenotype sex, removal of outliers who deviated +/- 3 standard deviations from the samples' heterozygosity rate mean as well as verification that the data did not contain closely related individuals (PI_HAT >0.2).

Prior to imputation, we determined the intersect of SNVs across three genotyping arrays, filtered each array for the uniform content (506,548 SNVs) and merged the data into one matrix. We used the Haplotype Reference Consortium reference panel[3] (webpage: http://www.haplotype-reference-consortium.org/) for array imputation with the following parameters: build hg19, reference panel apps@hrc-r1.1, population mixed, phasing eagle. After imputation, we filtered out SNVs with low imputation quality score R2 <0.3 and applied HWE <1e-6 and MAF <0.05 filters by European and African American ancestral group separately. For the final SNV set, we considered the union of filtered SNVs retrieved in both ancestry sets (8,245,179 biallelic SNVs). We tested for an association between imputed SNVs and genotyping array to confirm that SNV data were not biased towards any genotyping array. No bias towards any genotyping array was detected (**Supplementary Fig. 2**).

We used the 1000 Genome Project data[4] as reference to exclude samples that showed differential ancestry background than European or African/Admixed American based on the principal component analysis (PCA) (**Supplementary Fig. 3**).

RNA-sequencing data

Raw .fastq files (2 x 125bp pair-end reads) were downloaded from dbGaP. RNA-seq mapping was done with STAR 2-pass mapping[5] using GENCODE hg19 reference genome annotations. RNA-seq data quality was assessed using QTLtools bamstat mode[6] (webpage: https://qtltools.github.io/qtltools/) for i) the number of mapped sequencing reads passing mapping quality filter, ii) number of mapped sequencing reads falling within the GENCODE hg19 annotations, and iii) the number of GENCODE hg19 annotations covered by at least one sequencing read.

Gene expression was quantified by counting the number of RNA-seq reads that were in correct orientation, that had a mapping quality score above 225 and that did not contain more than 16 mismatches in both ends of the fragment with the reference genome. Each gene in each sample was normalized to get an RPKM value using the formula RPKM = (read_count x 1e9) / (total_mapped_reads x gene_length). We filtered for protein-coding and lincRNA genes and considered the union quantifications detected in >=50% in SCZ cases and in >=50% in controls. This yielded 18,258 protein-coding genes and 3,730 lincRNAs (total of 21,988 genes) for 243 individuals.

ChIP-sequencing data

Raw .fastq files (2 x 75bp pair-end reads; profiled for histone mark H3K27ac) were downloaded from dbGaP. Reads were aligned to reference genome hg19 with BWA-MEM v0.7.16[7] using an alignment score threshold of 10. For quality control, all ChIP-seq experiments were processed through Phantompeakqualtools v1.16[8] to generate two quality metrics: normalized strand cross-correlation (NSC) and relative strand cross-correlation (RSC) (**Supplementary Fig. 4**). These metrics use the cross-correlation of stranded read density profiles to measure enrichment independently of peak calling. RSC <0.8 and NSC <1.05 indicate low signal to noise ratio. The quality tag (based on thresholded RSC) was >2 (very high) for all experiments.

Peak calling and quantification were carried out with HOMER v14.11.1[9] (webpage: http://homer.ucsd.edu/homer/). Briefly, we first determined ChIP-seq peak coordinates across SCZ cases and controls to get a population scale call set of ChIP-seq peaks and then quantified the peaks for each individual according to the identified peak coordinates. Specifically, we first built a population call set of ChIP-seq peaks by aggregating 1e6 ChIP-seq reads from 74 SCZ cases and 74 controls together in a unique BAM file. Next, we carried out the actual peak calling onto the derived consensus peak set (derived unique BAM file) for all individuals (n=193: 74 SCZ cases and 119 controls) using HOMER findPeaks mode and parameters -style histone -o auto. This yielded 141,219 ChIP-seq peaks for each individual (mean peak length 2137 base pairs (bp)). Next, we quantified the peaks by obtaining per-peak read counts per sample using the peak coordinates from the consensus peak set and using the HOMER script annotatePeaks.pl with the following options: - noann -nogene -size given. This script counted the number of ChIP-seq reads falling within the peak coordinates. Read counts were subsequently normalized for a total of 10 million mapped reads per sample.

Mislabelling detection

Given that many analyses relied on testing genotype versus sequence data, we looked at the concordance between both to ensure there was no sample mislabelling and all individuals had both sequence and genotype data available. We used QTLtools mbv mode[6,10] and .vcf and .bam file for each individual and assessed the concordance at heterozygous and homozygous genotypes between genotype and RNA-sequencing and between genotype and ChIP-seq sequencing data. No such errors were detected.

COMPUTATIONAL AND STATISTICAL ANALYSES

Cis-regulatory domain (CRD) calling

For CRD calling, we used the pipeline developed in Delaneau et al. 2019[11] (webpage: https://github.com/odelaneau/clomics). We started with building a correlation map by systematically measuring interindividual correlation between all possible pairs of ChIP-seq peak quantifications (i.e., retrieved Pearson correlation coefficients using corrected and rank-normal transformed data matrix (see final paragraph of this section for specifics). Next, we applied an agglomerative hierarchical clustering of the data on a per chromosome basis. Specifically, we started with each ChIP-seq peak being assigned to its own cluster and iteratively merged clusters as we moved up in the hierarchy. To determine the pair of clusters to be merged, we maintained throughout the procedure a correlation matrix that corresponded to the matrix of squared correlations between ChIP-seq peaks, searched the pair of clusters that exhibited the highest value as well as constantly updated the correlation matrix when merging clusters together. For minimizing computational cost, we searched for the maximal correlation between clusters that were not separated by more than 250 rows or columns (i.e., peaks or groups of peaks in the correlation matrix), allowing to store and update the diagonal parts of the correlation matrix and merging together proximal features of the genome. This strategy resulted in a binary tree that regrouped all ChIP-seq peaks from the same chromosome in which each node delimited a set of highly correlated ChIP-seq peaks.

CRDs were called by identifying the minimal set of internal nodes that captured most of the overall correlation mass (i.e., cumulative sum of squared correlation). To retain an internal node as a CRD, three criteria needed to be fulfilled: i) CRDs regrouped only highly correlated ChIP-seq peaks: the mean absolute correlation between all possible pairs of ChIP-seq peaks within a CRD had to be at least twice as high as the mean correlation between all ChIP-seq peaks in the chromosome; ii) CRDs had well-defined boundaries: the mean absolute correlation between all pairs of ChIP-seq peaks involving either the first or the last ChIP-seq peak (on the basis of their genomic location) had to be at least twice as high as the same value derived for the first and last peaks on the chromosome; iii) CRDs captured distal coordination between at least two regulatory elements (REs): ChIP-seq peaks had to cover at least two non-overlapping regulatory regions. These three criteria were implemented into an algorithm that processed each binary tree starting from the root node (node regrouping all peaks of a chromosome) and recursively traversed the internal nodes of the tree until an internal node fulfilled all three criteria. Then, declared the internal node and all the peaks downstream as a CRD, stopped to go deeper by ignoring the children of this node and carried on with other internal nodes in the tree.

This pipeline was applied using only SCZ ChIP-seq quantifications (n=74), only control ChIP-seq quantifications (n=119) and using ChIP-seq quantifications in the combined set (i.e., SCZ cases and controls together; n=193), such that we started with three separate correlation maps and in the end had called SCZ-specific CRDs, controls-specific CRDs as well as uniform CRDs across all samples. For correcting ChIP-seq quantifications, we identified the optimal number of PCs that captured variability in ChIP-seq data in SCZ cases, in controls and across samples via QTL mapping (see section QTL mapping for molecular phenotypes) by considering variable number of PCs as covariates. After finding the optimal configuration (giving the best QTL discovery power; **Supplementary Fig. 5a-c**), we corrected ChIP-seq peak quantifications for 3 genotype PCs and 10 ChIP-seq PCs in SCZ cases (n=74), 3 genotype PCs and 20 ChIP-seq PCs in controls (n=119), and 3 genotype PCs and 30 ChIP-seq PCs across samples (n=193). All three resulting data matrices were rank-normal transformed separately.

CRD activity quantification

For CRD activity quantification, we applied a dimensionality reduction approach, i.e., we enumerated all ChIP-seq peaks per CRD, and took the mean of all single peak quantifications per individual to retrieve a single quantification value for each individual. We used the ChiP-seq peak correlation map retrieved in the combined set (i.e., across SCZ cases and controls; n=11,374 CRDs). For SCZ-specific CRD structure analyses, we retrieved CRD activity quantifications for SCZ cases only using the CRDs identified in SCZ cases (n=10,938 CRDs). The resulting vectors were again rank-normalized such that these matched a normal distribution with mean 0 and standard deviation 1 and consisted of one row per CRD and one column per individual.

Quantitative Trait Loci (QTL) mapping for molecular phenotypes (ChIP-seq peak activity, gene expression, CRD activity)

For each molecular phenotype, we first enumerated all genetic variants within +/- 1 Mb and then tested each one of these variants for association with the phenotype and only retained the best hit (i.e., with the smallest nominal p-value). Secondly, we adjusted the best nominal p-value for the number of variants being tested by permutations. Specifically, we randomly shuffled the phenotype quantifications 1,000 times and retained the best association p-values for each permuted data set, which effectively gave 1,000 null p-values of associations. Third, to correct for the number of molecular phenotypes being tested whole genome (e.g., number of genes, peaks, CRDs), we used a false discovery rate (FDR) correction approach and declared phenotype-variant pairs at FDR 5% threshold as significant. These steps were carried out with QTLtools cis mode[6].

To discover multiple QTLs with independent effects on a given molecular phenotype, we used the conditional analysis approach implemented in QTLtools[6]. Briefly, this approach is based on a forward-backward scan of the cis-window around the phenotypes to automatically learn the number of independent QTLs and to identify the most likely candidate variants, while controlling for a given FDR.

For SCZ-specific QTL discovery, we considered QTL effects identified in SCZ cases (796 aCRD-QTLs and 867 eQTLs) and for each variant-phenotype pair ran a linear regression including genotype, disease status (SCZ/CTL), and covariates, and tested for significance of a genotype * disease status interaction on molecular phenotype (gene expression or CRD activity). This was followed with FDR 5% correction for the number of QTLs tested.

CRD structure analysis

For determining CRD sharing between SCZ cases and controls, we compared ChIP-seq peak correlation maps between SCZ cases and controls and called a CRD shared if ≥50% of the peaks overlapped between the reference and the query correlation map.

To assess the features of SCZ-specific CRDs, we considered only CRDs in SCZ cases composed of peaks not regrouping into any CRD in controls. These formed 28% of the CRDs identified in SCZ cases (3,078 CRDs composed of 6,650 peaks). For underlying peak activity estimation, we used ChIP-seq peak quantifications normalized for 10 million reads per sample, uncorrected for any covariates and applied a Mann-Whitney U test per peak activity between SCZ cases and controls. Significant differences between SCZ cases and controls were determined at FDR 5% using *R/qvalue* package[12].

For confirming whether the peaks within SCZ-specific CRDs showed different correlation structures in SCZ cases vs controls and were not driven by the mean background correlation estimate ascertained separately in SCZ cases and controls in CRD calling, we used ChIP-seq peak quantifications corrected for biological and technical covariates (3 genotype PCs and 10 ChIP-seq PCs in SCZ cases, and 3 genotype PCs and 20 ChIP-seq PCs in controls as outlined in CRD calling section). The corrected data matrices were rank-normal transformed separately. We calculated the mean Pearson correlation estimate between peak activities per CRD separately in SCZ cases and controls (i.e., in controls measured the correlation estimate between peaks per SCZ-specific CRDs) and used Mann-Whitney U test for comparing identified mean correlation estimates between SCZ cases and controls.

CRD and gene association

Briefly, we considered normalized CRD activity quantifications (final step in CRD activity quantification section) and corrected and normalized gene expression quantifications and used QTLtools cis permuation pass[6] to identify CRDs associated with a gene in a +/-1 Mb window from a gene's transcription start site. We performed these analyses to i) identify genes associated with SCZ-specific CRDs, and ii) capture comparable associations for SCZ cases and controls using the same CRD annotations, i.e., CRDs identified in the combined set. The first approach was performed in SCZ cases only (n=59) using CRD activity quantifications retrieved based on the CRDs identified in SCZ cases (10,938 CRDs; **Supplementary**

**Table 1)**. The second approach was done for SCZ cases (n=59), for controls (n=105) and across samples (n=164; disease status (SCZ/CTL) considered as a covariate) using CRD activity quantifications retrieved based on the CRDs identified in the combined set (**Supplementary Table 5**).

Specifically, to capture technical and biological variability in gene expression data, we residualized for ancestry, using 3 genotype PCs, and for the number of optimal RNA-seq PCs that allowed to discover the maximum number of eQTLs. This was done similarly as for the ChIP-seq data by doing association testing at variable number of PCs. Gene expression quantifications were corrected for 3 genotype PCs and 10 RNA-seq PCs in SCZ cases, 3 genotype PCs and 30 RNA-seq PCs in controls, and 3 genotype PCs and 40 RNA-seq PCs in the combined set (**Supplementary Fig. 5d-f**). The resulting matrices were rank-normal transformed. Next, we enumerated all CRDs within +/-1 Mb of gene's transcription start site, tested their activity for association with gene expression and stored the best hit together with the nominal p-value. We adjusted the nominal p-value for the number of CRDs being tested in cis using permutation and corrected for the number of genes being tested using the R/*qvalue* package[12]. We determined gene-CRD associations at FDR 5% as significant.

Differential CRD activity and differential gene expression analysis

Both differential CRD activity and differential gene expression analyses were carried out using DESeq2[13]. Significant associations were determined at FDR 5% (**Supplementary Table 2, Supplementary Table 3**). For differential CRD activity analysis, we used unnormalized ChiP-seq peak read counts obtained with HOMER (annotatePeaks.pl with options -noann -nogene -size given -raw)[9] and summed these up per CRD using the ChiP-seq peak correlation map identified in the combined set (11,374 CRDs). For differential gene expression analysis, we used RNA-seq read counts.

To identify covariates for correction, we carried out association testing i) between all available biological and technical covariates and diagnosis status (Mann-Whitney U test), and ii) between all available biological and technical covariates and individual ChIP-seq peak activity and gene expression quantifications (linear regression) and calculated $\pi 1$ estimate[12] to identify the proportion of true associations. We identified the following covariates for differential CRD activity analysis: sex, age at death, genotype PC1, library batch, GC content in sequencing data, empirical insert size, 15bp repeat in sequencing data. We identified the following covariates for differential gene expression analysis: sex, age at death, genotype PC1, genotype PC2, post-mortem interval, brain pH, brain weight, RNA integrity number, total RNA yield, A260/A280 ratio, GC content in sequencing data, transcript integrity number, empirical insert size, 15bp repeat in sequencing data, date of sequencing.

R*/clusterProfiler* package[14] was used for gene set enrichment analysis. We considered genes that were either i) significantly differentially expressed (regardless of direction of effect), ii) significantly down-regulated or iii) significantly up-regulated. Significant associations were determined at FDR 5% (**Supplementary Table 4**).

Association of aCRD-QTLs and eQTLs with the other molecular phenotype

We tested QTL effects for association with the other molecular phenotype (i.e., gene expression with aCRD-QTLs and CRD activity with eQTLs) via CRD-gene nominal associations using CRD identified in the combined set). Specifically, for each gene we identified in cis window all associated CRDs at nominal pass, and for each CRD determined in cis window all associated genes at nominal pass. Using these intermediate associations, we could look whether e.g., an eQTL also affects the CRD that the targeted eGene is associated with, and vice versa, whether the aCRD-QTL affects the gene that the impacted aCRD is associated with. Proportion of sharing was estimated using $\pi 1$ estimate[12].

Colocalization with SCZ GWAS variants

Briefly, we assessed the likelihood of a shared functional effect between SCZ risk variants from four GWAS studies[15-18] and SCZ-identified QTLs (796 aCRD-QTLs and 876 eQTLs) using regulatory trait concordance (RTC). Specifically, we considered independent hits from four GWAS studies and applied the RTC algorithm. This algorithm assesses the likelihood of a shared functional effect between a GWAS variant and a QTL variant by quantifying the change in the statistical significance of the QTL after correcting the

QTL phenotype (gene expression or CRD activity) for the genetic effect of the GWAS variant and comparing its correction impact to that of all other SNPs in the interval[19,20]. We applied a cut-off of RTC $\geq$0.9 for determining a shared functional effect. The output files indicate the union results across four GWAS studies. When accounting for LD, we see 16 independent shared effects between GWAS and eQTL variants and 12 independent shared effects between GWAS and aCRD-QTL variants (**Supplementary Table 9**).

CRD-gene pair quantification

To quantify the 1,197 gene-CRD pairs we identified as significant at FDR 5% across SCZ cases and controls (n=164), we used PCA-based dimensionality reduction. For each gene-CRD pair, we aggregated gene expression with CRD activity and used the coordinates on PC1 as new pseudo-phenotypes. For identifying genetic variants that affect both the CRD activity and the gene expression (per gene-CRD pair), we used the new derived pseudo-phenotypes and carried out an eCRD-QTL (genetic variant that affects both the CRD and gene) discovery analysis in cis across all samples (n=164) using permutation.

Causal inference for determining causal relationships for eQTLCRD-CRD-gene triplets

We applied a Bayesian Network approach to infer the most likely causal relationship for eCRDQTL-CRD-gene triplets common to SCZ cases and controls (1,100 triplets) and conducted the analyses separately in SCZ cases (n=59) and in controls (n=105). This approach allowed to estimate the most likely network from which the observed data originates. The starting point is always the genetic variant as this does not change (genome is fixed). We explored three distinct models (topologies): i) causal model in which the genetic variant affects first the CRD and then the gene, ii) reactive model in which the genetic variant affects the gene and then the CRD, iii) independent model in which the genetic variant affects the gene and the CRD independently (**Supplementary Fig. 17**). For each triplet we built a 59 x 3 and 105 x 3 data matrix for SCZ cases and controls, respectively, that contained normalized quantifications, and calculated the likelihood of three possible Bayesian Network topologies using R/*bnlearn* package[21]. We converted the likelihoods to posterior probabilities, assuming a uniform prior probability for three possible models.

Bootstrapping

To estimate the accuracy for the Bayesian Network results and provide confidence for retrieved probabilities, we used bootstrapping. We carried out 100 bootstrapping runs for each tested triplet separately for SCZ cases (n=59) and controls (n=105) using sampling with replacement. For accuracy estimation, we calculated how many times the most probable model across bootstrapping runs for each triplet was the same as in the original Bayesian Network results. We filtered out all triplets that fell below a confidence threshold of 55%: this corresponds to the lower quartile value in SCZ cases (**Supplementary Fig. 19ab**).

Gene set enrichment for model-change associated triplets

Used R/*clusterProfiler* package[14] for gene set enrichment analysis for genes that belonged to triplets showing directional change from eCRD-QTL onto gene expression/CRD activity between SCZ cases and controls. We considered two scenarios: i) causal model in controls, but reactive/independent in SCZ cases (n=147); ii) reactive/independent in controls, but causal in SCZ cases (n=87).

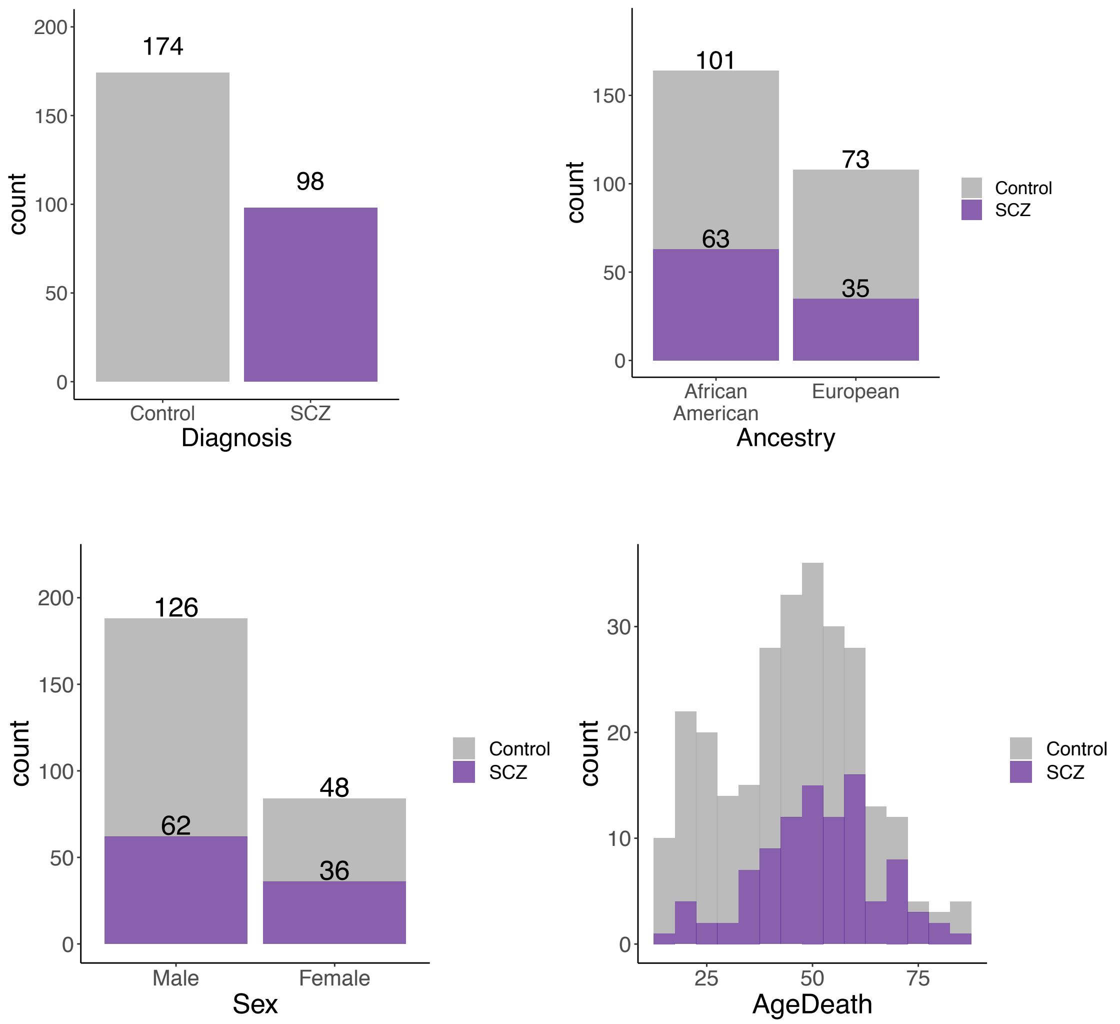


**Supplementary Fig. 1. Overview of the phenotypic characteristics of the HBCC cohort**. Mean age 51 years (sd = 14.4) for schizophrenia (SCZ) cases and 42.2 years (sd = 16.4) for controls.

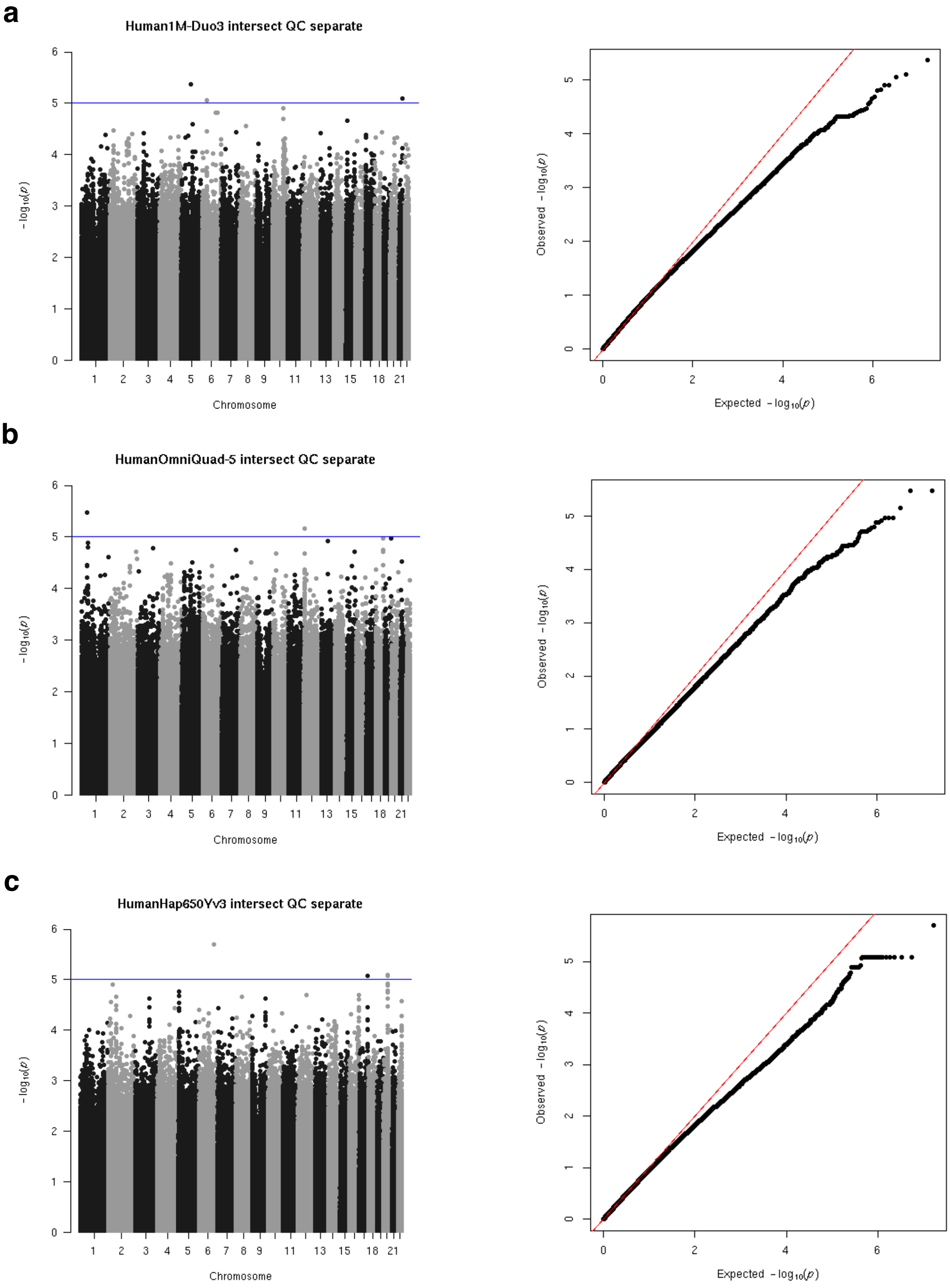


**Supplementary Fig. 2. Association testing between imputed SNVs and genotyping array.** For each genotyping array, an association test was carried out using logistic regression for array of interest (either (a) Illumina Human1M-Duov3, (b) Illumina HumanOmni5M-Quad or (c) Illumina HumanHap650Y) vs other two arrays. Manhattan and QQ-plots depict the distribution of imputed SNVs per chromosome as a function of -log10 p-values and expected vs observed p-values, respectively. Post-imputation quality control (MAF <0.05 and HWE <1e-6) was applied by ancestry and the union SNV content across ancestry sets was considered for this and all downstream analyses. No bias towards any genotyping arrays was detected.

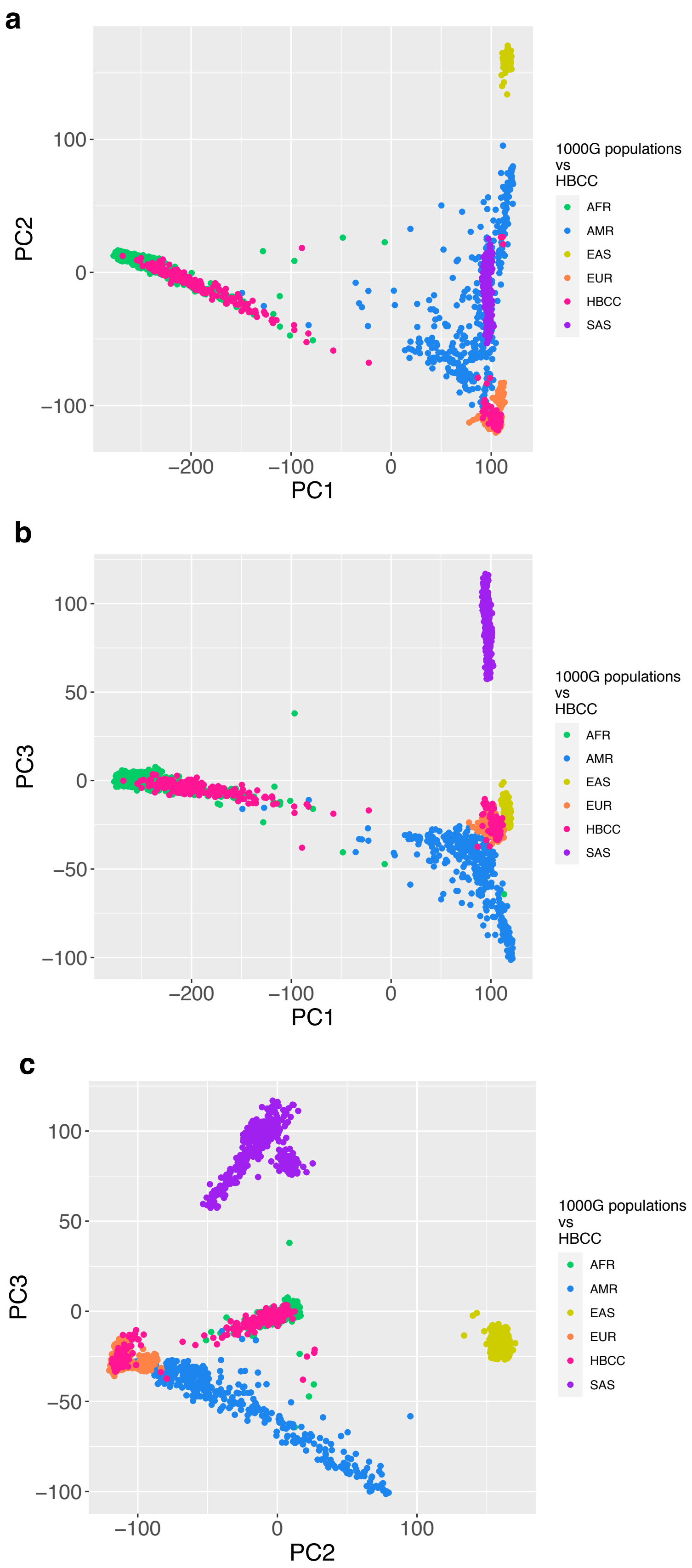


**Supplementary Fig. 3. Principal component (PC) analysis of genotype data.** Coordinates of (a) PC1 vs PC2, (b) PC1 vs PC3 and (c) PC2 vs PC3 in reference to the 1000 Genome Project samples. HBCC cohort samples are coloured in pink and cluster at PC coordinates represented by the African and European 1000 Genome Project super populations.

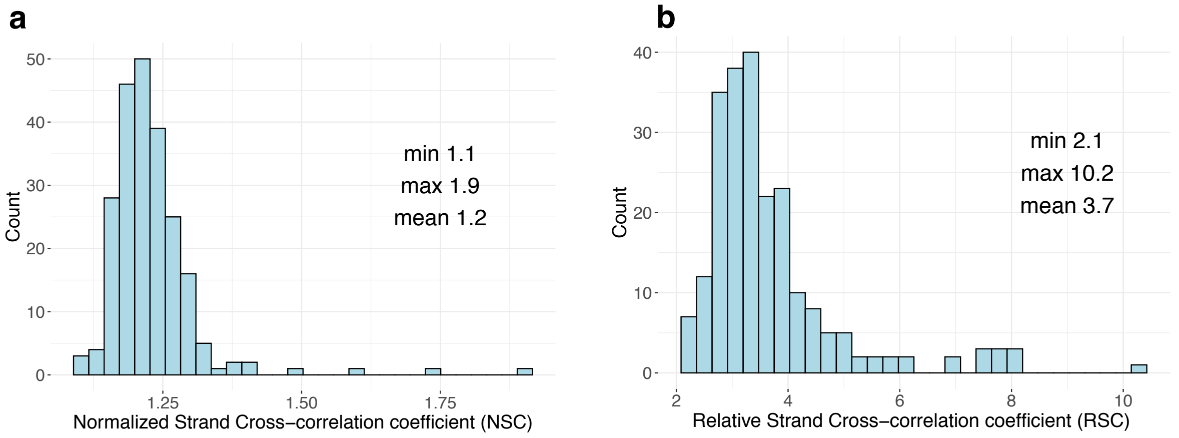


**Supplementary Fig. 4. PhantomPeakQualTools quality metrics**. (a) Normalized Strand Cross-Correlation coefficient (NSC) distribution and (b) Relative Strand Cross-correlation coefficient (RSC) distribution for H3K27ac peaks in HBCC samples. NSC <1.05 and RSC <0.8 indicate low signal to noise.

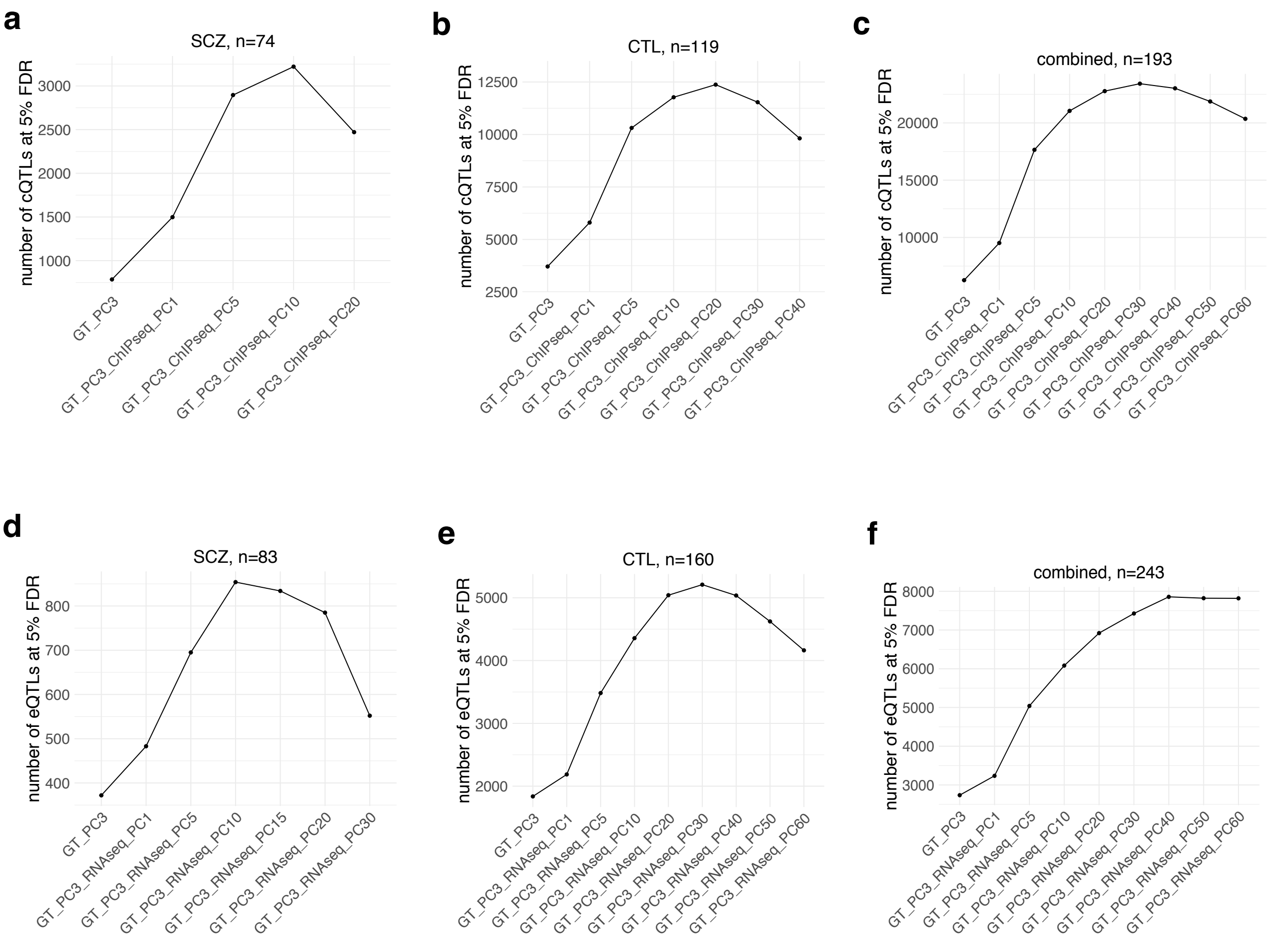


**Supplementary Fig. 5. Optimization of QTL discovery.** Number of chromatin-QTLs (cQTLs) and expression-QTLs (eQTLs) discovered (a,d) in SCZ cases, (b,e) in controls (CTL) and (c,f) in the combined set as a function of the number of genotype (GT) principal components (PCs) and ChIP-seq and RNA-seq PCs used to residualize ChIP-seq peak quantification and gene expression quantification data, respectively. The PCs that allowed the discovery of the maximum number of QTLs were retrained for downstream analyses.

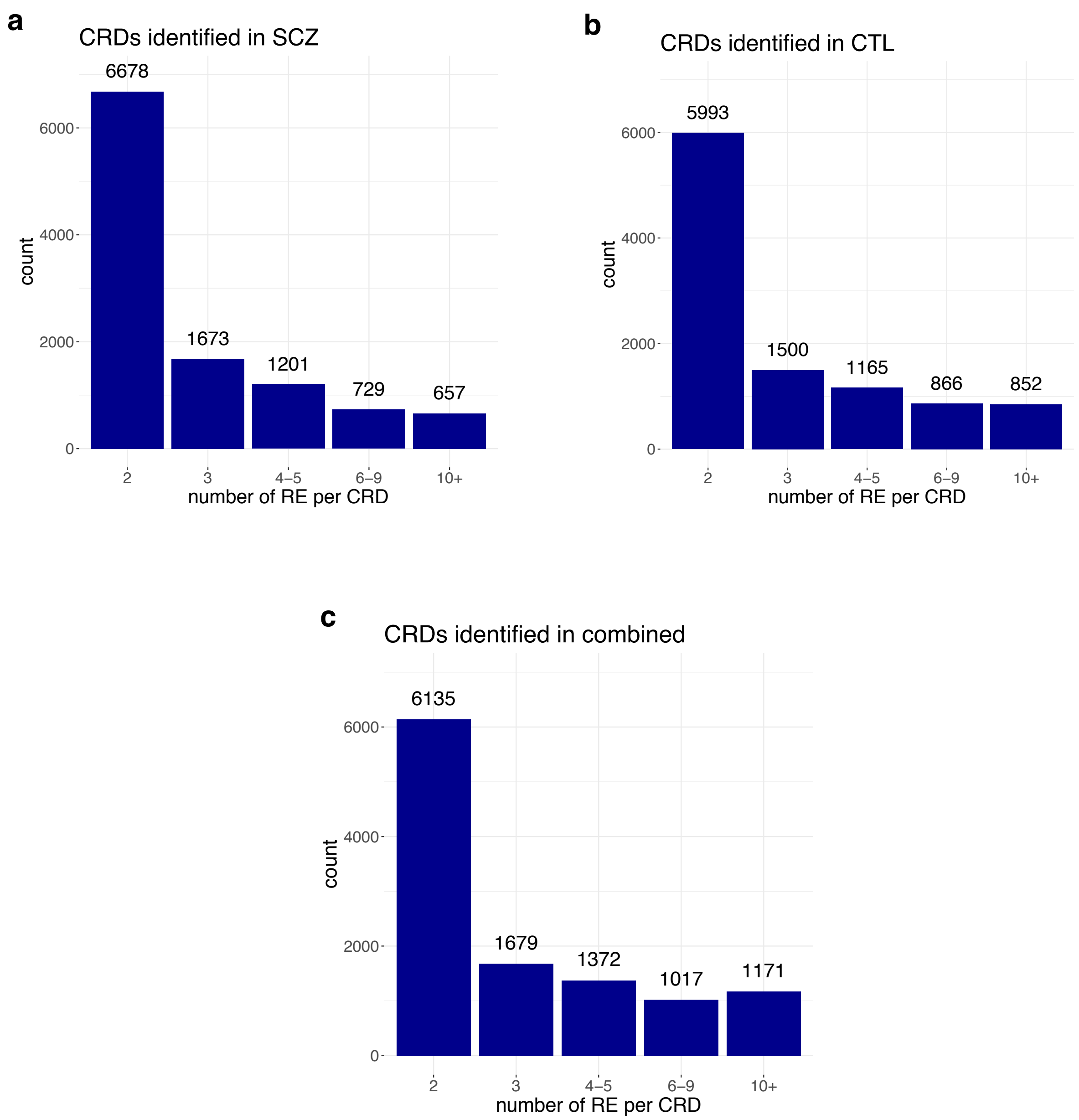


**Supplementary Fig. 6. Regulatory element (RE) content of CRDs.** Number of regulatory elements per CRD (a) in SCZ cases (n=74), (b) in controls (CTL, n=119), and (c) in the combined set (n=193). Mean number of REs per CRD was 3.7, 4.3 and 4.7 in SCZ cases, in controls and in the combined set, respectively. Mean CRD length was 137,017 bp, 135,734 bp and 138,144 base pairs in SCZ cases, in controls and in the combined set, respectively.

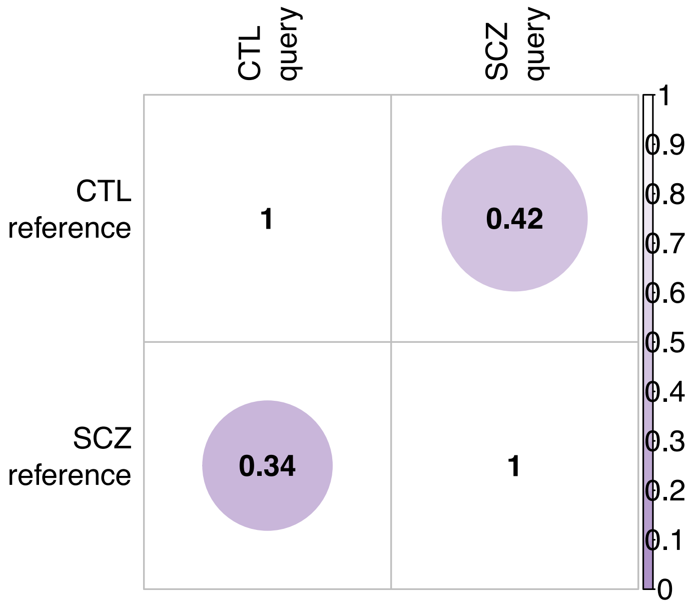


**Supplementary Fig. 7. Fraction of CRD peak content sharing between SCZ cases and controls (CTL).** A CRD was deemed shared between SCZ cases and controls in case ≥50% of ChIP-seq peaks of the reference CRD were present in a CRD from the query state. Forty-two percent of CRDs detected in controls (n=119) were also detected in SCZ cases (n=74) and vice versa, 34% of CRDs detected in SCZ cases were also detected in controls.

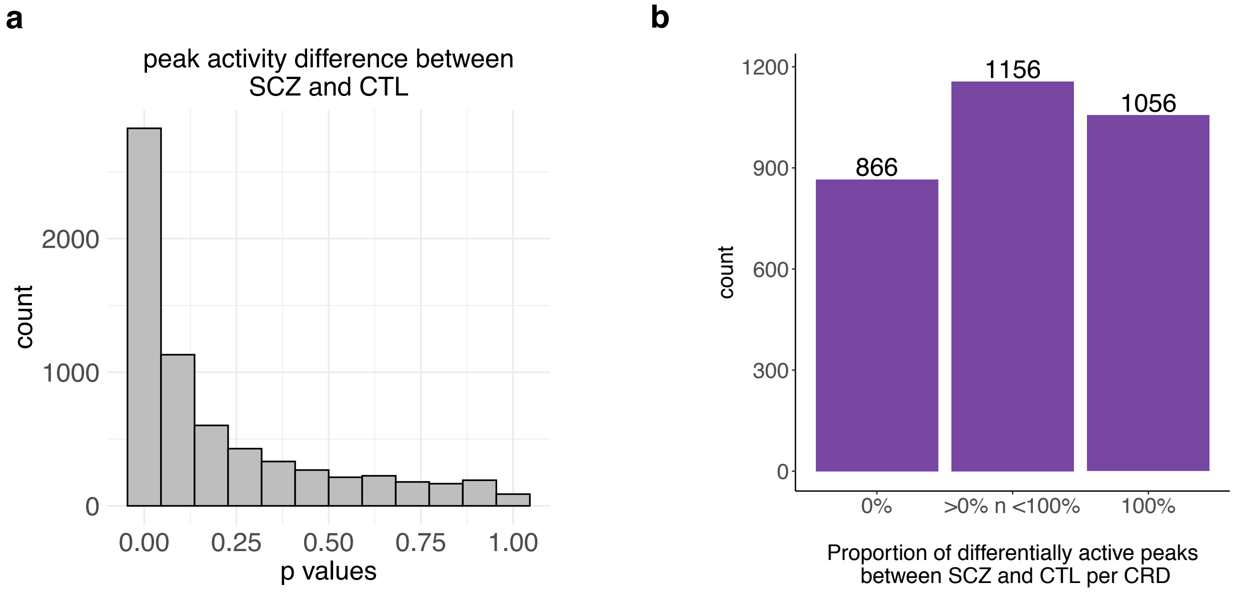


**Supplementary Fig. 8. Differential peak activity between SCZ cases and controls (CTL)**. Only peaks clustering into SCZ-specific CRDs (i.e., CRDs composed of peaks not part of any CRD in controls) were considered. (a) P-value distribution of peaks differentially active between SCZ cases and controls. (b) Proportion of differentially active peaks (3,540 peaks) between SCZ cases and controls per SCZ-specific CRDs at FDR 5%. One-third of SCZ-specific CRDs (1,056 CRDs and 2,242 peaks) were forming due to all underlying peaks showing significantly different peak activity in SCZ cases compared to controls.

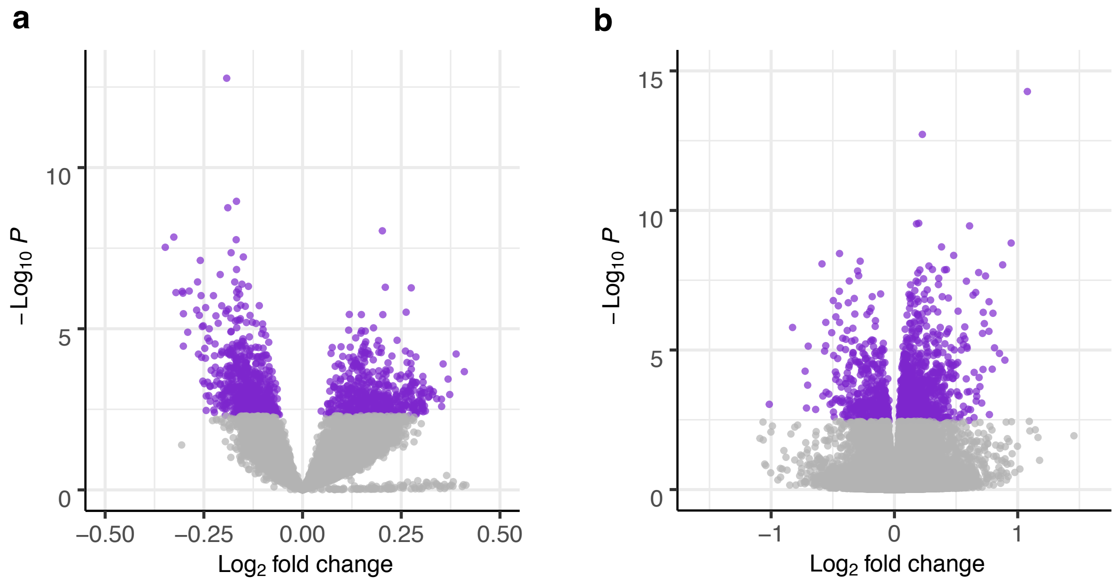


**Supplementary Fig. 9. Volcano plots outlining (a) differential CRD activity and (b) differential gene expression between SCZ cases and controls.** Differentially active CRDs (n=1,141; 599 with lower activity and 542 with higher activity) and differentially expressed genes (n=1,363; 937 up-regulated and 426 down-regulated), respectively, at FDR 5% are highlighted in purple.

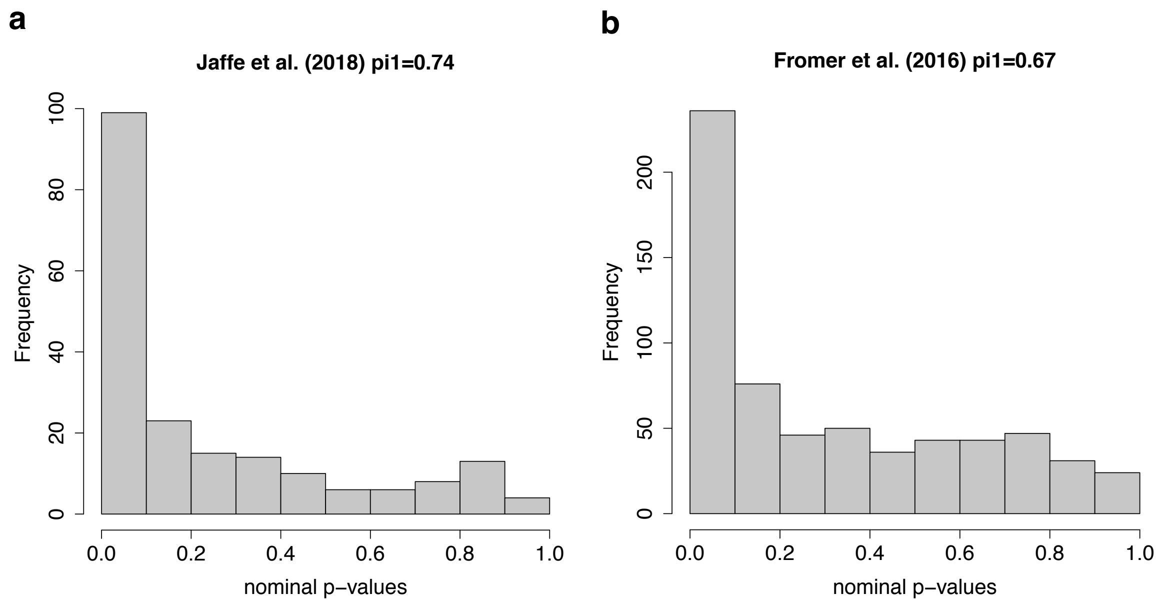


**Supplementary Fig. 10. Replication of differential gene expression analysis results for SCZ in previously published findings.** Differentially expressed genes identified at FDR 5% were in concordance with findings published by (a) Jaffe et al., 2018 and by (b) Fromer et al., 2016 based on π1 estimate.

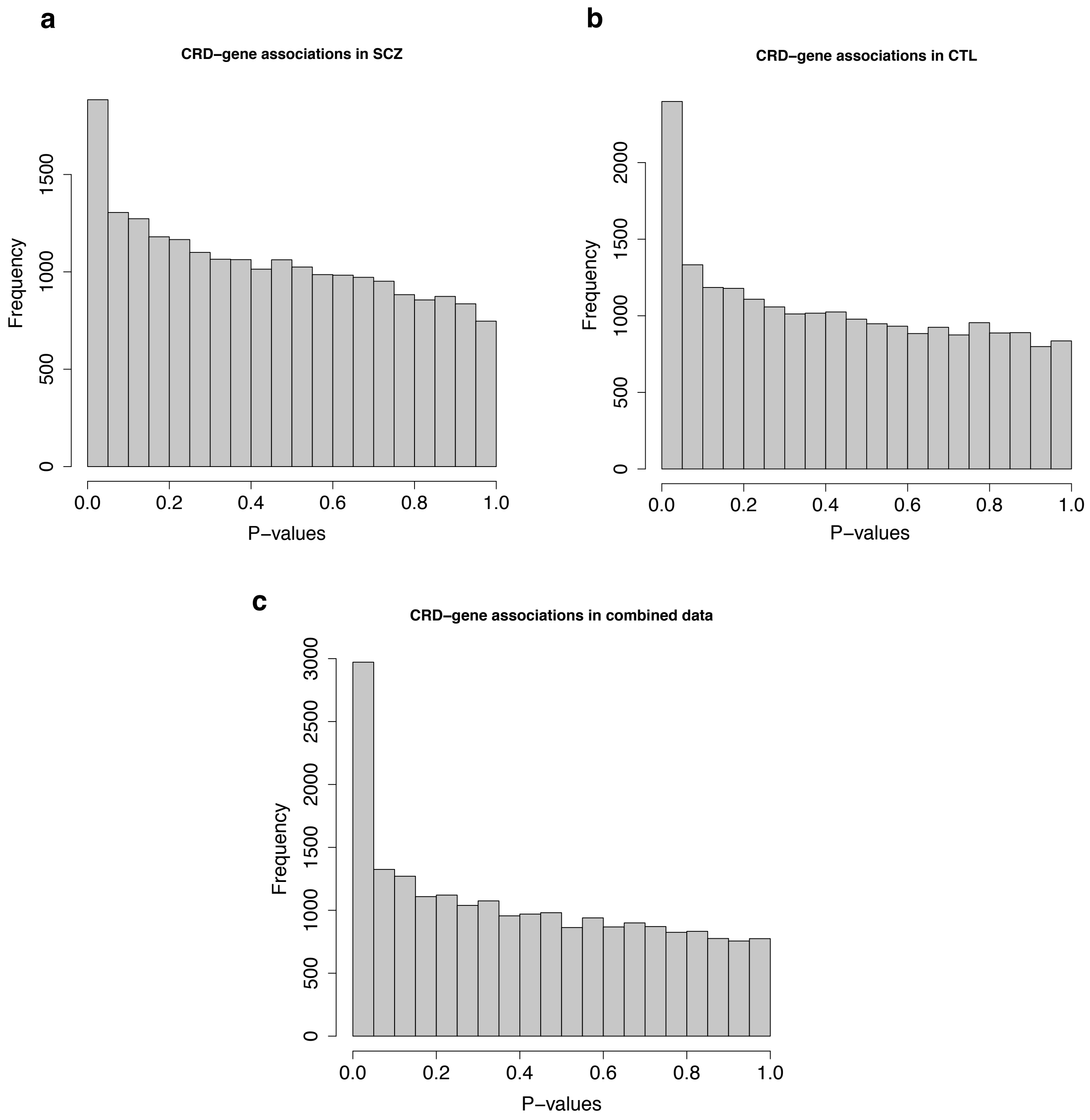


**Supplementary Fig. 11. Gene-CRD associations.** P-value distribution for gene-CRD associations (a) in SCZ cases (n=59), (b) in controls (n=105) and (c) in the combined set (n=164). At FDR 5%, 95, 634 and 1,197 CRD-gene associations were identified in SCZ cases, in controls and in the combined set, respectively. Disease status (SCZ/CTL) was considered as a covariate for CRD-gene pair detection across in the combined set.

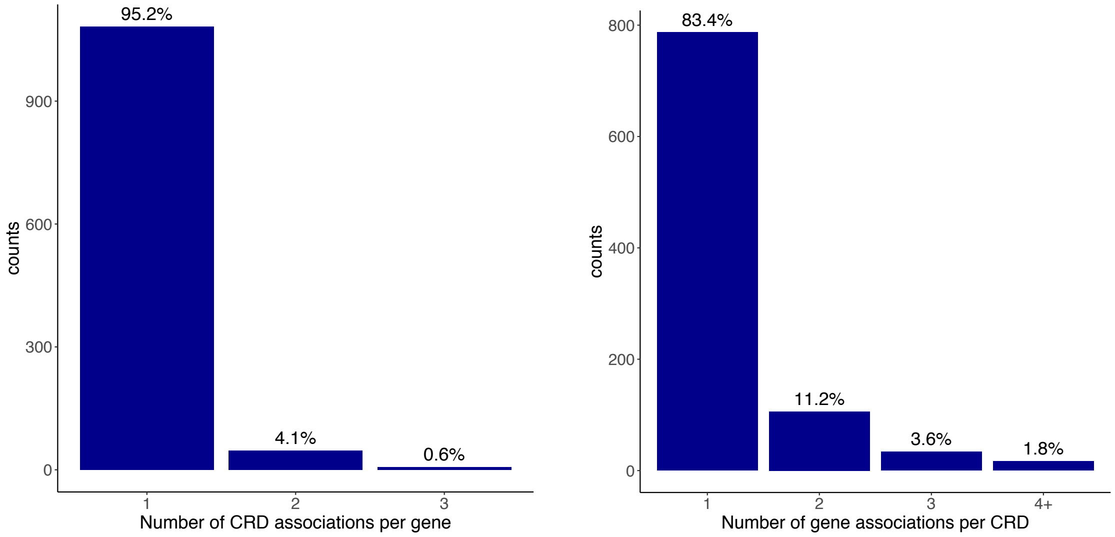


**Supplementary Fig. 12. Gene-CRD associations.** Number of genes and CRDs as a function of the number of CRDs and genes they were associated with, respectively, identified in the combined set (n=1,197 gene-CRD associations).

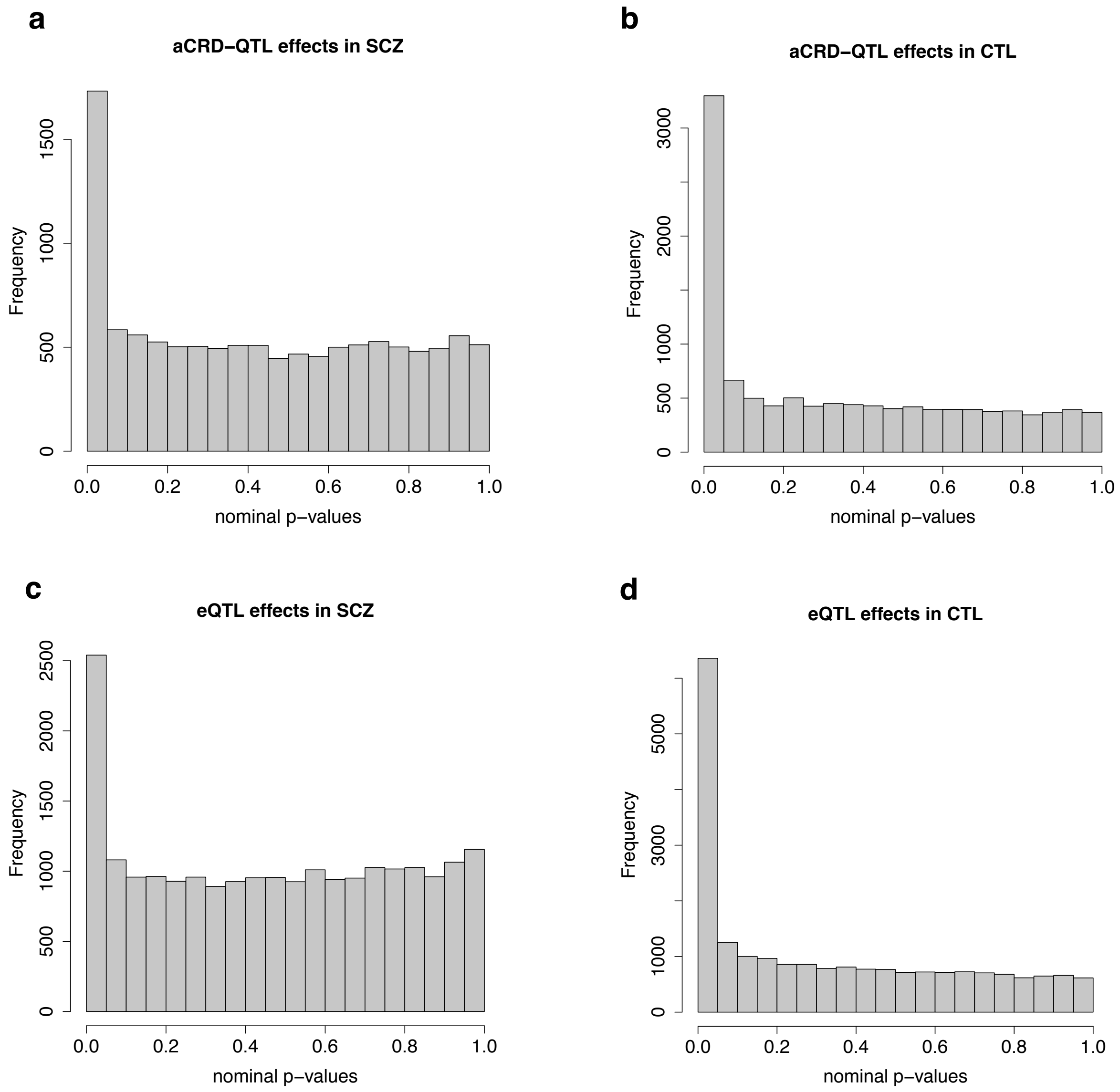


**Supplementary Fig. 13. QTL discovery.** P-value distribution for QTL and CRD activity/gene expression associations (a,c) in SCZ cases and (b,d) in controls. At 5% FDR and in cis, 796 and 2,929 functionally independent aCRD-QTLs, and 867 and 6,166 functionally independent eQTLs in SCZ cases and controls, respectively, were discovered.

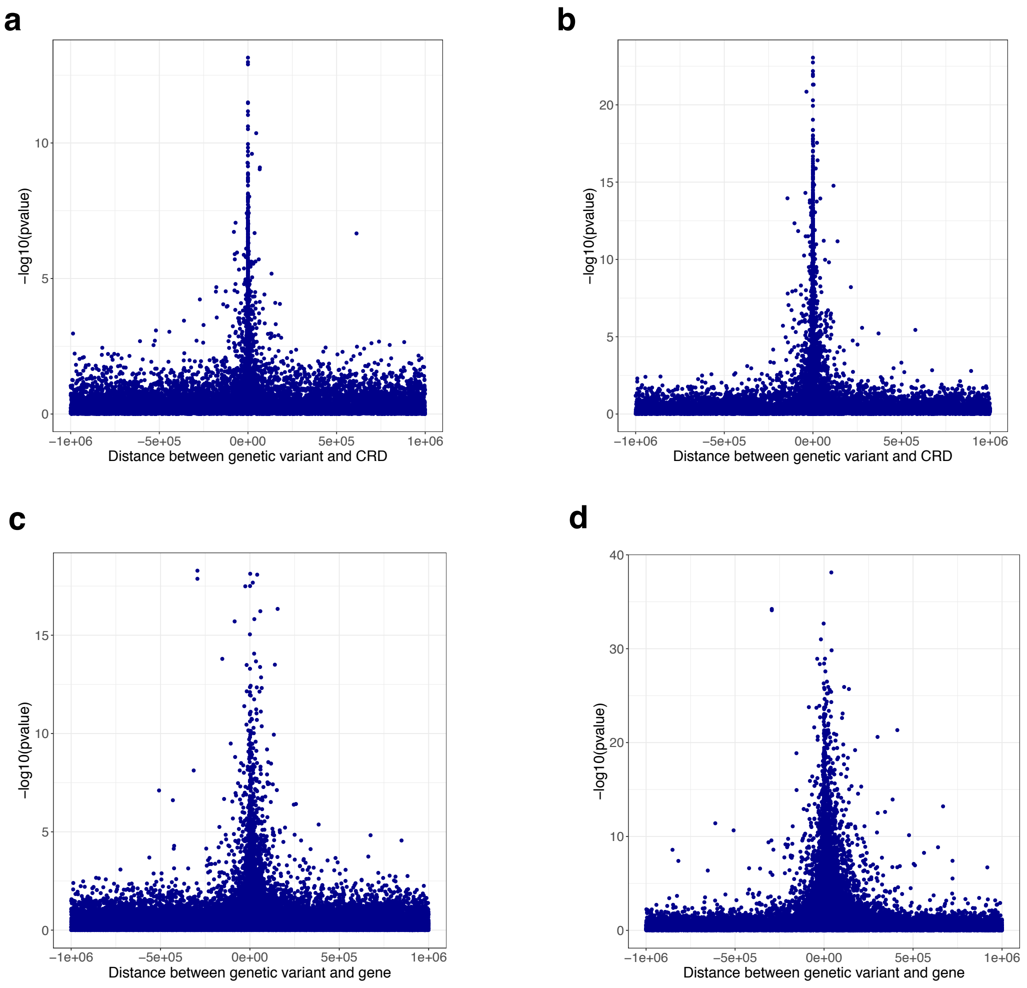


**Supplementary Fig. 14. CRD/gene distance in base pairs from associated QTL.** Genomic distance between genetic variant and CRD/gene as a function of the strength of association given in -log10 p-values (a,c) for SCZ cases and (b,d) for controls.

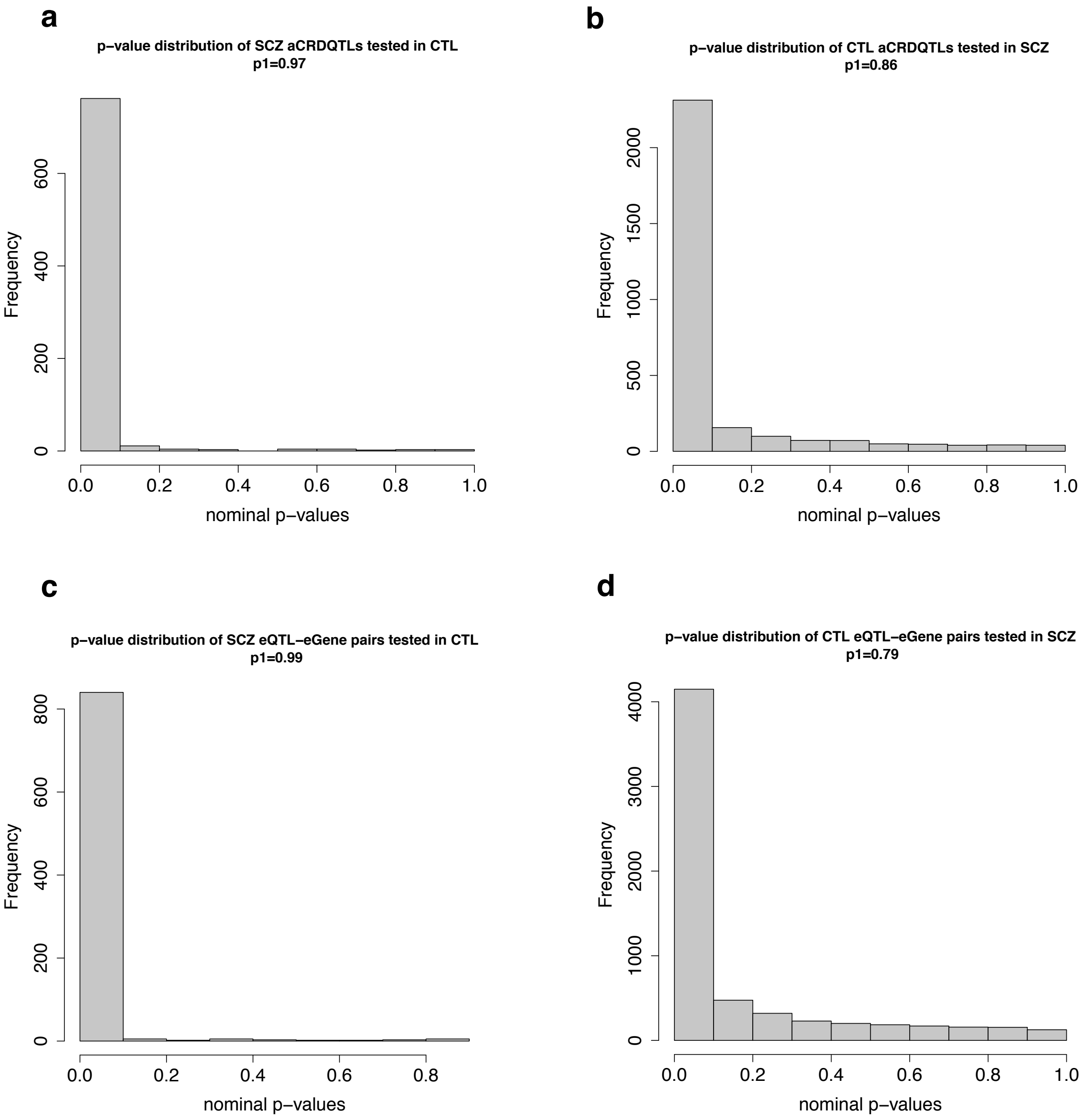


**Supplementary Fig. 15. Proportion of sharing aCRD-QTL and eQTL effects between SCZ cases and controls based on π1 estimate.** P-value distributions of SCZ-identified (a) aCRD-QTL and (c) eQTL effects tested in controls, and control-identified (b) aCRD-QTL and (d) eQTL effects tested in SCZ cases.

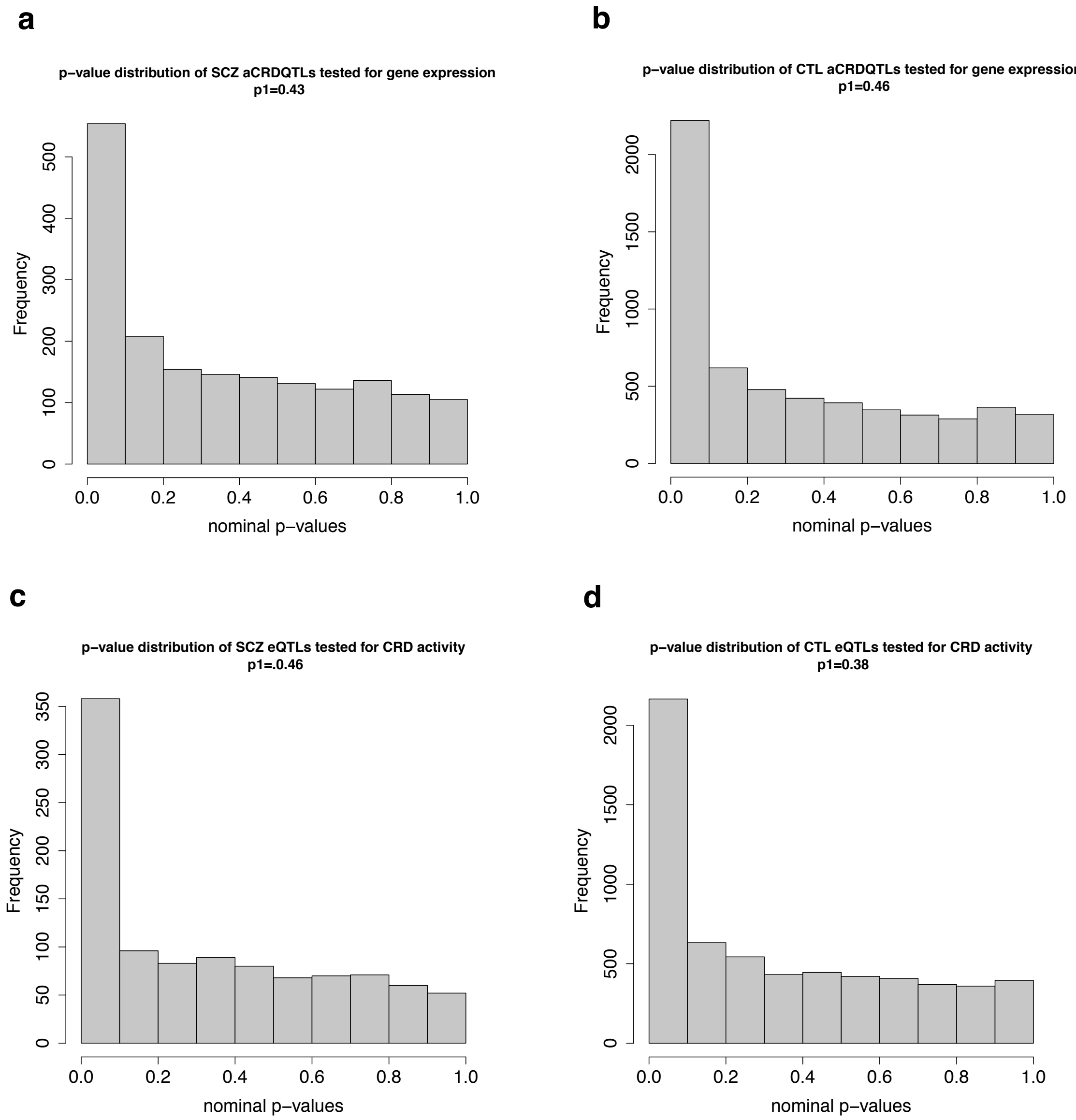


**Supplementary Fig. 16. Proportion of sharing aCRD-QTL and eQTL effects for gene expression and for CRD activity, respectively, based on π1 estimate.** P-value distribution of (a) SCZ-identified and (b) controls-identified aCRD-QTLs tested for gene expression over CRD-gene associations identified at nominal significance level. P-value distribution of (c) SCZ-identified and (d) controls-identified eQTLs tested for CRD activity over gene-CRD associations identified at nominal significance level.

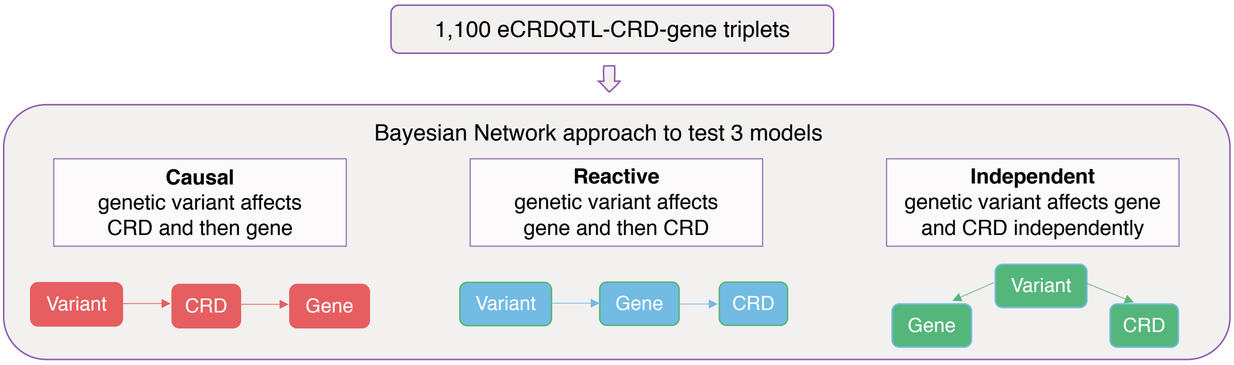


**Supplementary Fig. 17. Bayesian Network approach.** Schematic of models considered in Bayesian Networks to infer the most likely causal relationship for eCRDQTL-CRD-gene triplets in SCZ cases and controls. eCRD-QTL denotes a genetic variant that affects the activity of a CRD and the expression of a gene that show a significant association with each other.

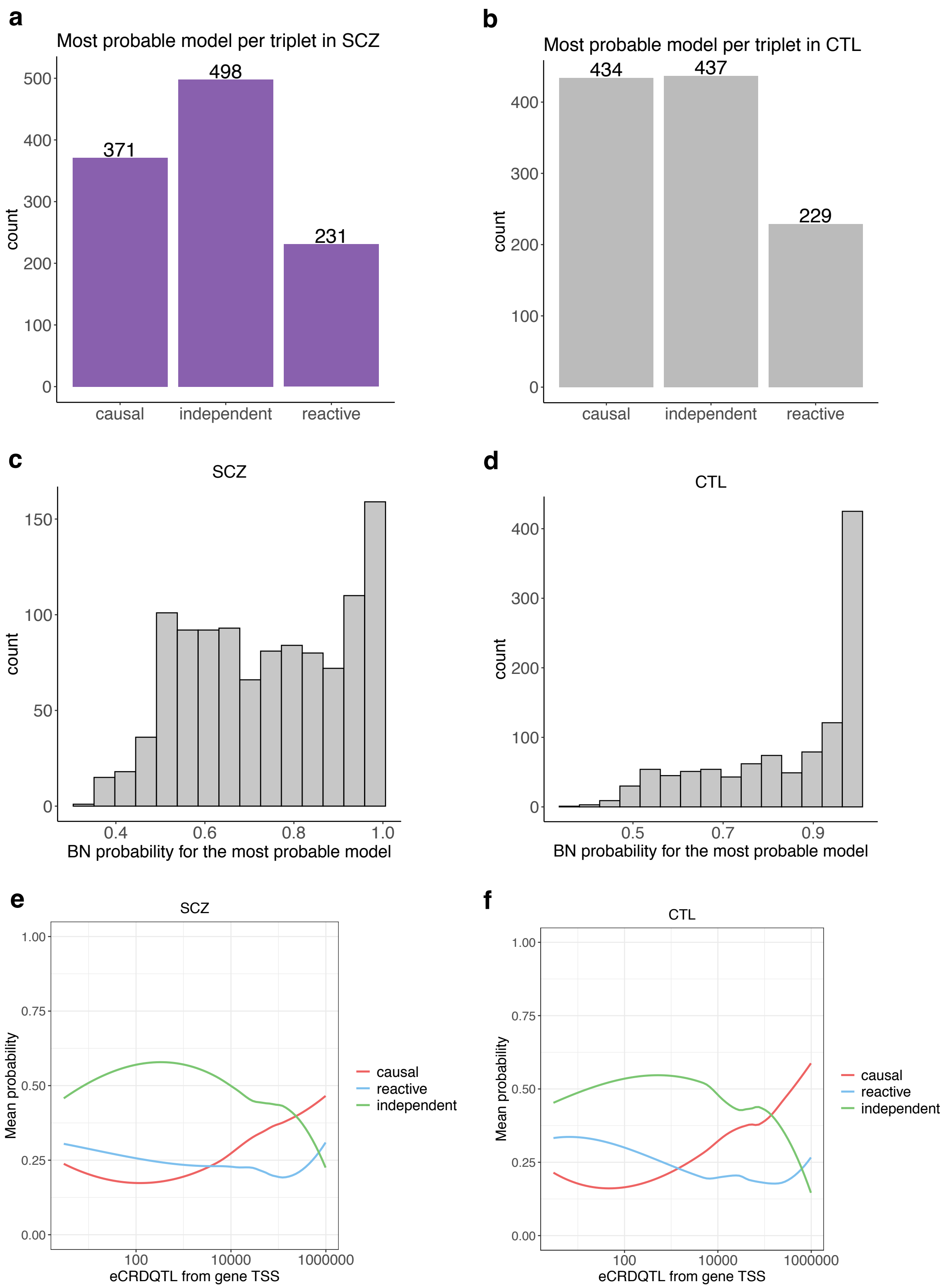


**Supplementary Fig. 18. Causal inference estimation for eCRDQTL-CRD-gene triplets.** Counts of the most probable model for each triplet (n=1,100) for (a) SCZ cases and (b) controls. Distribution of the probabilities for the most probable model for each triplet for (c) SCZ cases and (d) controls. Mean probabilites for each model as a function of the distance in base pairs between eCRD-QTL and the target gene for (e) SCZ cases and (f) for controls.

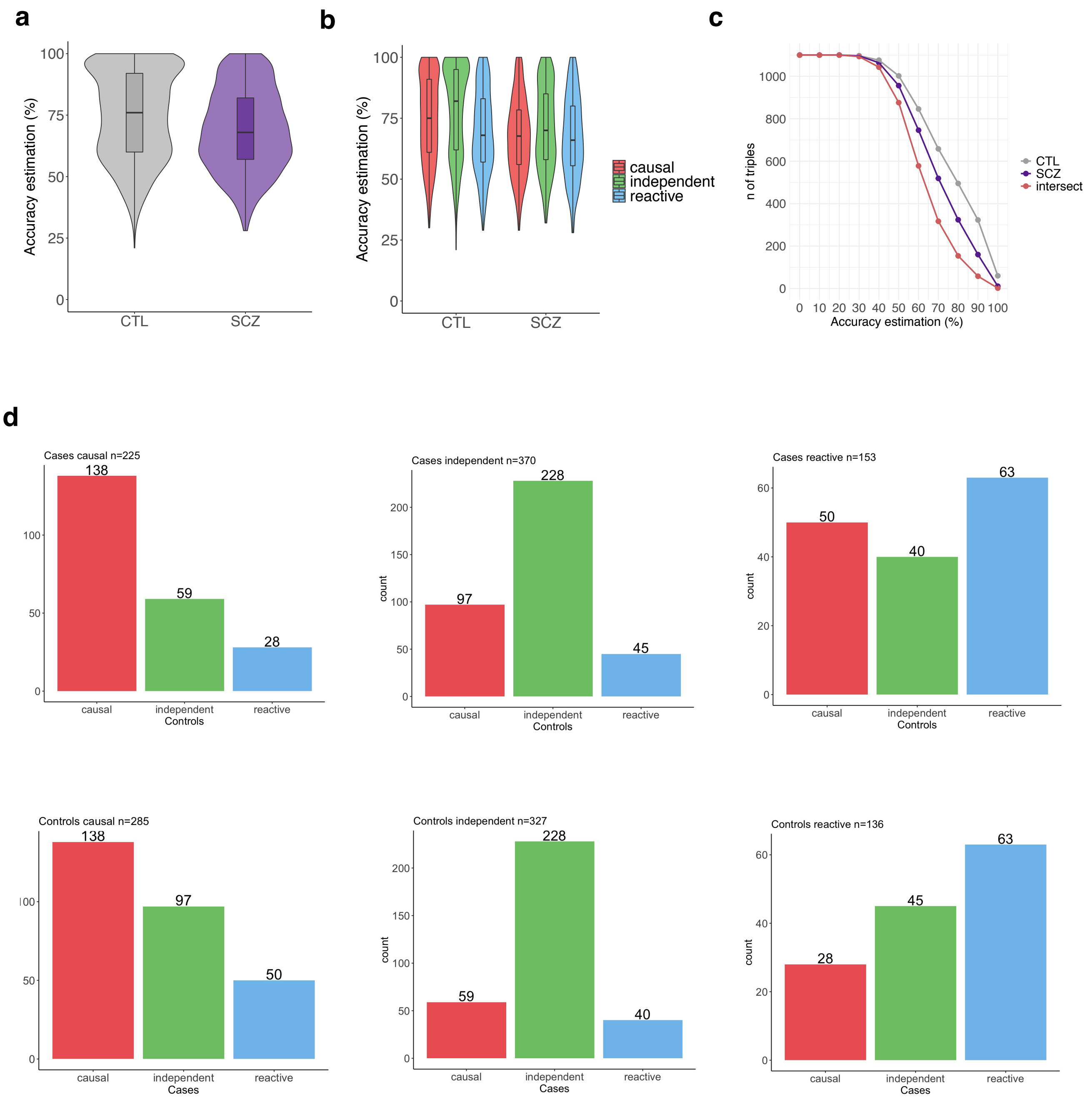


**Supplementary Fig. 19. Regulatory mechanism for eCRDQTL-CRD-gene triplets in SCZ cases and controls.** Distribution of accuracy estimation (%) denoting how often the most probable model across bootstrapping runs for each triplet was the same as in the original Bayesian Network analysis for SCZ cases and controls (a) across models and (b) by model. (c) Triplet counts as a function of accuracy estimation; purple colour denotes triplet counts for SCZ cases, grey for controls and red indicates triplet counts at the intersect of accuracy estimation for SCZ cases and controls. (d) Comparison of the direction of effect from eQTL-CRD onto molecular phenotypes between SCZ cases and controls for 748 triplets that surpassed the accuracy estimation of 55%.

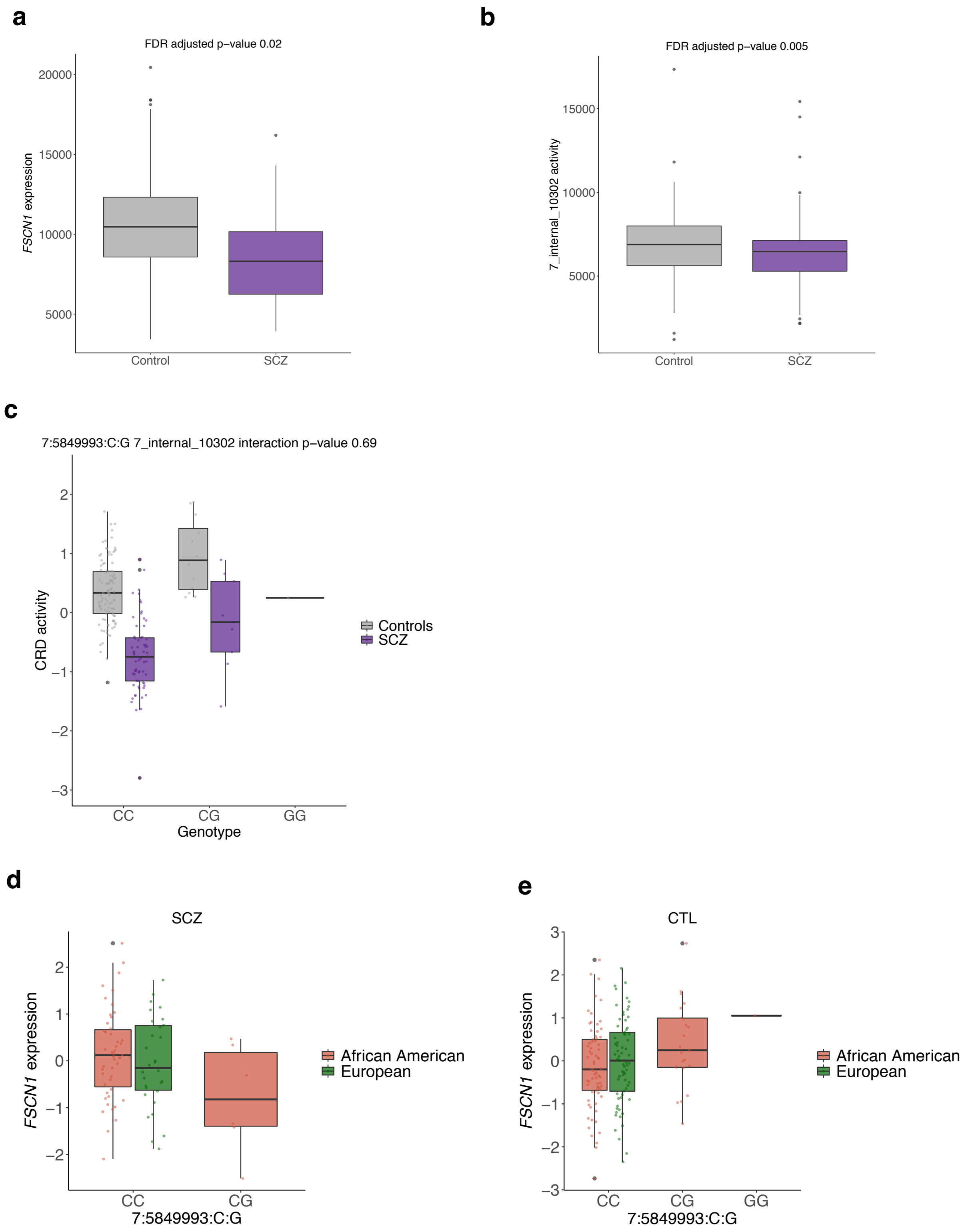


**Supplementary Fig. 20. Example of mechanistic change in the regulation of gene expression between SCZ cases and controls for a triplet consisting of an eCRD-QTL 7:584993:C:G, gene *FSCN1* and a CRD composed of 5 REs on chr7:5623132-5705414.** Distribution of (a) *FSCN1* gene expression and (b) CRD activity for SCZ cases and controls. (c) Genotype-dependent effect for eCRD-QTL 7:584993:C:G on CRD activity. Genotype-dependent effect for eCRD-QTL 7:584993:C:G on *FSCN1* expression by ancestry group (d) for SCZ cases and (e) for controls.
